# Colder and drier winter conditions are associated with greater SARS-CoV-2 transmission: a regional study of the first epidemic wave in north-west hemisphere countries

**DOI:** 10.1101/2021.01.26.21250475

**Authors:** Jordi Landier, Juliette Paireau, Stanislas Rebaudet, Eva Legendre, Laurent Lehot, Arnaud Fontanet, Simon Cauchemez, Jean Gaudart

**Affiliations:** IRD, Aix Marseille Univ, INSERM, SESSTIM, Marseille, France; Mathematical Modelling of Infectious Diseases Unit, Institut Pasteur, UMR2000, CNRS, Paris, France; Santé publique France, French National Public Health Agency, Saint Maurice, France; Hôpital Européen Marseille, France; Emerging Infectious Diseases Unit, Institut Pasteur, Paris, France; PACRI Unit, Conservatoire National des Arts et Métiers, Paris, France; Aix Marseille Univ, APHM, INSERM, IRD, SESSTIM, Hop Timone, BioSTIC, Marseille, France

## Abstract

Higher transmissibility of SARS-CoV-2 in cold and dry weather conditions has been hypothesized since the onset of the COVID-19 pandemic but the level of epidemiological evidence remains low.

During the first wave of the pandemic, Spain, Italy, France, Portugal, Canada and USA presented an early spread, a heavy COVID-19 burden, and low initial public health response until lockdowns. In a context when testing was limited, we calculated the basic reproduction number (R0) in 63 regions from the growth in regional death counts. After adjusting for population density, early spread of the epidemic, and age structure, temperature and humidity were negatively associated to SARS-CoV-2 transmissibility. A reduction of mean absolute humidity by 1g/m3 was associated with a 0.15-unit increase of R0. Below 10°C, a temperature reduction of 1°C was associated with a 0.16-unit increase of R0.

Our results confirm a dependency of SARS-CoV-2 transmissibility to weather conditions in the absence of control measures during the first wave. The transition from summer-to winter-like conditions likely contributed to the intensification of the second wave in north-west hemisphere countries. Adjustments of the levels of social mobility restrictions need to account for increased SARS-CoV-2 transmissibility in winter conditions.

## Introduction

The spread of SARS-CoV-2 in February-March 2020 caught the majority of European and North American countries unprepared. The spread of the virus was largely uncontrolled until movement restriction and social distancing policies were put in place at national or regional level with various degrees of intensity [1]. Most regions that implemented lockdowns experienced a peak in hospital admissions approximately 2 weeks after the lockdown was imposed, corresponding to infections acquired around the date of lockdown [2].

The spread of this first wave was largely heterogeneous between countries. Various determinants were proposed to explain the differential spread of the virus. Some Asian countries like Japan, South Korea, Vietnam and Thailand and some isolated countries like Australia, New Zealand, Iceland, experienced only limited transmission and stood out for high levels of preparedness and efficient response strategies. Even within South West European and North American countries where no effective response was deployed before lockdowns, SARS-CoV-2 virus spread heterogeneously at the regional level, as observed from hospitalization, death counts, and confirmed by serological surveys [3, 4].

Epidemic spread is characterized by the basic reproduction number, or R0. R0 expresses the average number of secondary cases resulting from a given case in the context of a naive population.

R0 depends on individual susceptibility to infection when in contact with an infective individual, and on the probability of an infectious contact. Environmental parameters affect R0: population density may increase the probability of contacts between individuals and weather conditions may affect the survival of the virus or the individual susceptibility to an infection. Most known respiratory viruses spread during the cold season in the temperate Northern hemisphere [5]. Weather conditions in winter can also affect individual susceptibility to infection through irritation of the nasal mucosa, but also influence the behaviour of individuals towards conditions prone to transmission (living or gathering in closed, heated spaces with a dry atmosphere) [5]. In addition, temperature, humidity, and UV, might directly affect the virus survival and modify infectiousness [6, 7]. Individual preventive behaviours (masks), collective strategies reducing mobility and contacts (lockdowns) or limiting the duration of the infectious period (detection and isolation) modify the number of secondary cases and the effective reproduction number can be calculated, which accounts for these alterations in the “natural” history of transmission.

In spite of a large number of studies, the evidence regarding the link between weather conditions and SARS-CoV-2 transmission remains limited. At date of 15 May 2020, a systematic review retained 61 studies analysing the relationship between COVID-19 epidemic and environmental factors [8]. Methodological issues included the lack of controlling for confounding factors such as population density [8]. Inappropriate epidemiological and statistical methods were also pointed out [8, 9]. Comparison between countries with different counter epidemic responses, testing strategy, or delayed onset of the epidemic might also have led to inconsistent results [8]. An earlier review retaining 17 studies highlighted the risk of bias and the low level of available evidence [10]. Between 15 May 2020 and 15 December 2020, our systematic search identified 82 research articles, of which only 15 (18%) analysed the growth rate or the reproduction number of SARS-CoV-2 (Appendix p2). Only four studies of climate and reproduction number included adjustments for confounding factors, and three of four included relevant covariates of population mobility, population density, and took into account interventions when necessary [11–13]. Of these, two were conducted over small geographical units: one study of 212 US counties identified a negative relationship between temperature increase and SARS-CoV-2 growth, while the other study including 28 Japanese prefectures identified a positive relationship [11, 12]. The third study was at the global scale for 203 states (in the USA, Canada, Australia and China) or countries and identified a negative association between UV light exposure and SARS-CoV-2 growth. Temperature was negatively associated with growth only after adjustment for UV light exposure [13]. All three studies identified a positive association between population density and epidemic growth.

Overall, multiple studies described a negative relationship between temperature and COVID-19 outcomes but the majority were unadjusted ecological correlation studies with strong risk of bias bringing low quality evidence [8], and evidence from multivariable growth studies remains ambiguous.

A modelling study defined the range of possible dependency between SARS-CoV-2 transmission and absolute humidity, based on two known coronaviruses and influenza [14], and recent models still do not include specific SARS-CoV-2 data [15]. More precise estimations of the effect of meteorological conditions on the spread of SARS-CoV-2 are required to better anticipate and inform policies regarding seasonal adjustments [16].

The objective of this study was to evaluate the contribution of weather parameters in the transmission of SARS-CoV-2, by analysing their effect on SARS-CoV-2 basic reproduction number in a context of low public health response during the early phase of the first wave in 6 north-western hemisphere countries.

## Results

### Region selection

The six countries (USA, Canada, Spain, Italy, France and Portugal) included 128 regions/states. Overseas regions (n=11) and regions which had experienced <10 cumulative deaths 28 days after lockdown (n=15) were not included (Figure 1). Likewise, regions with a maximal daily mortality <5 deaths (n=19) within 28 days after lockdown were not included. Overall, 83 regions were assessed for exponential growth period, and R0 was calculated for 64 regions with sufficient exponential growth (Figure S1 in Appendix): 24 regions in the USA, 2 regions in Canada, 11 regions in France, 12 regions in Spain, 13 regions in Italy and 1 region in Portugal.

**Figure 1:**
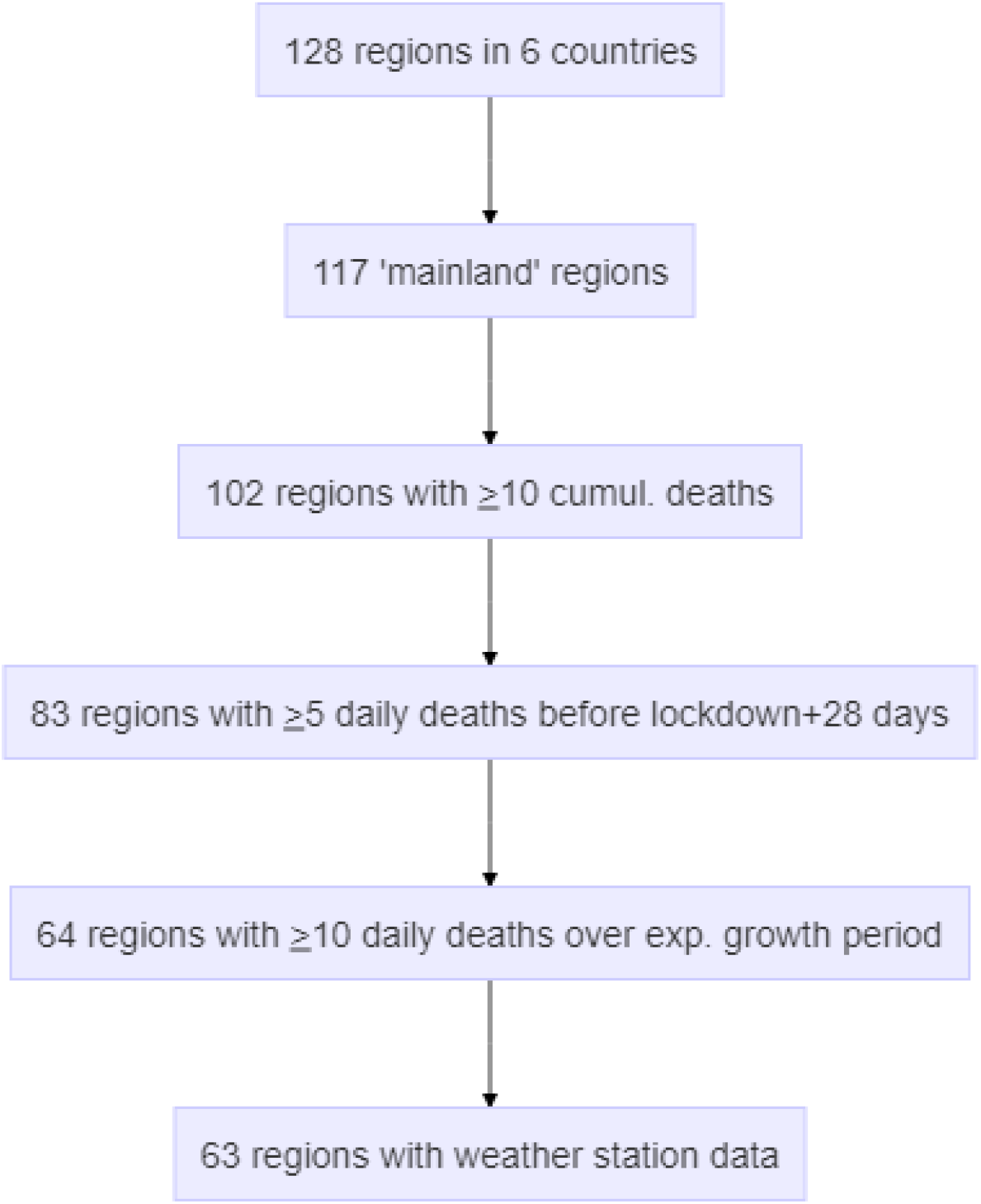
Study flow chart.

### Data description

The maximal daily death count was 39 deaths/day in median (interquartile range (IQR)=18-83, max=779) and occurred 25 days (median; IQR=19-43) after the start of the lockdown. Of note, larger reductions in human mobility patterns after lockdown as assessed from google mobility led to shorter delay (Spearman correlation coefficient = 0.63, p<0.0001, Figure S3). The delay between the start of lockdown and the peak in daily death count was reduced to 19 days (median; IQR=17-27) for South European countries with nationwide lockdowns and strong mobility reductions (>60% in average, Figure S3).

R0 was estimated over an exponential growth period that had a median duration of 11 days (IQR=9-14, range=5-19). In The R0 estimation period started 5 days (median; IQR=0-8.5) and ended 16 days after the date of lockdown (IQR=11-21, range 1-27). Figure S4 presents details of the calculation periods.

The median R0 value was 2.58 (IQR=2.08-2.66). R0 estimates were lowest (<1.5) in two regions in France and one in Spain, the USA and Italy (respectively in Centre-Val de Loire, Nouvelle Aquitaine, La Rioja, Alabama and Abruzzo). R0 was highest (>4.0) in New York (USA), Lombardia and Piemonte (Italy), Castilla-La Mancha (Spain) and Ontario (Canada)(Figure 3A and B). Spain and Canada had overall higher R0 values compared to Italy, France and USA (Figure 4A). R0 values exhibited significant spatial autocorrelation (Moran’s I=0.20, p=0.0087).

**Figure 2:**
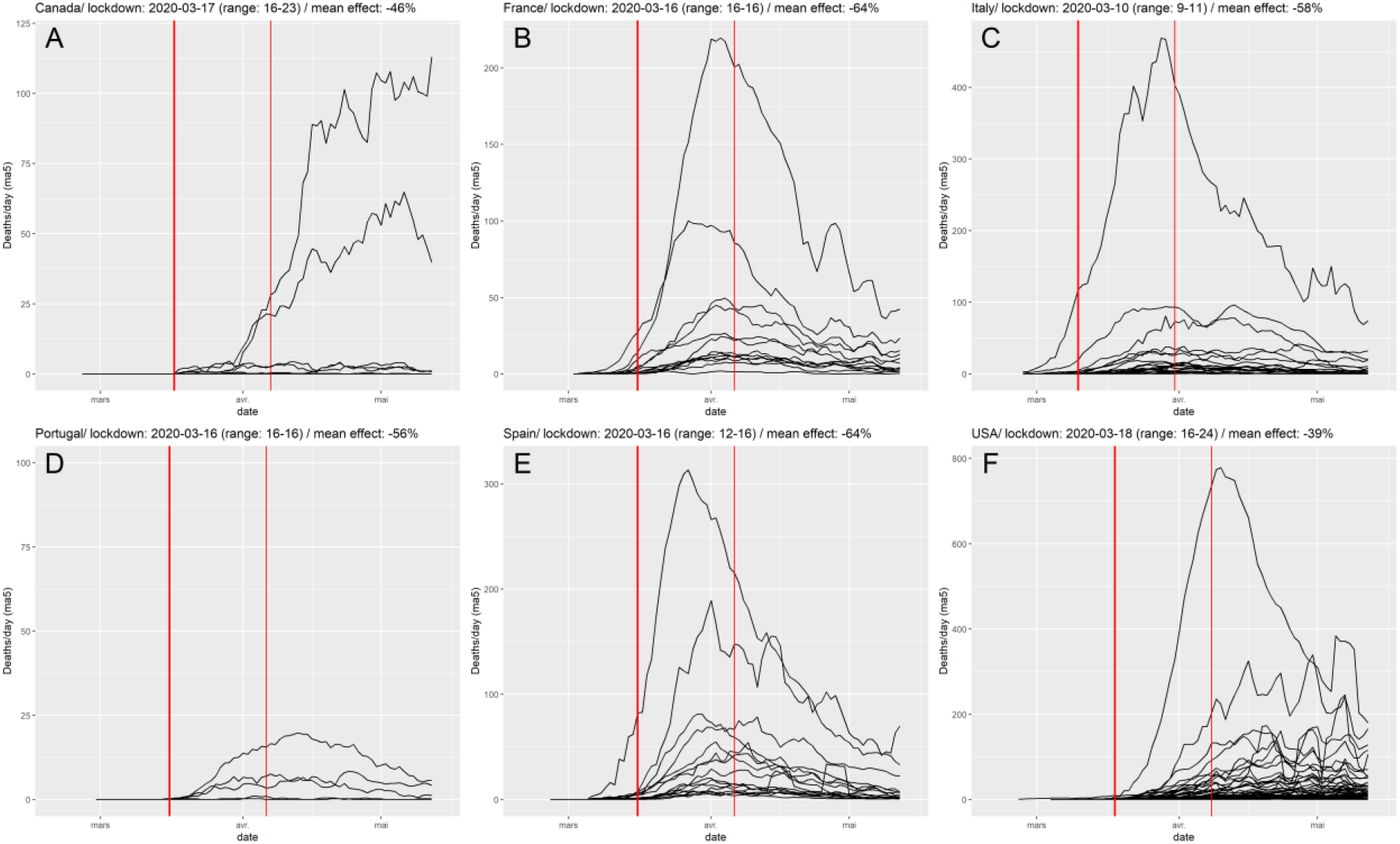
Deaths per day, by region and by country. The thick red line figures the median date of lockdown by each country, and the thin red line the median date +28 days.s

**Figure 3:**
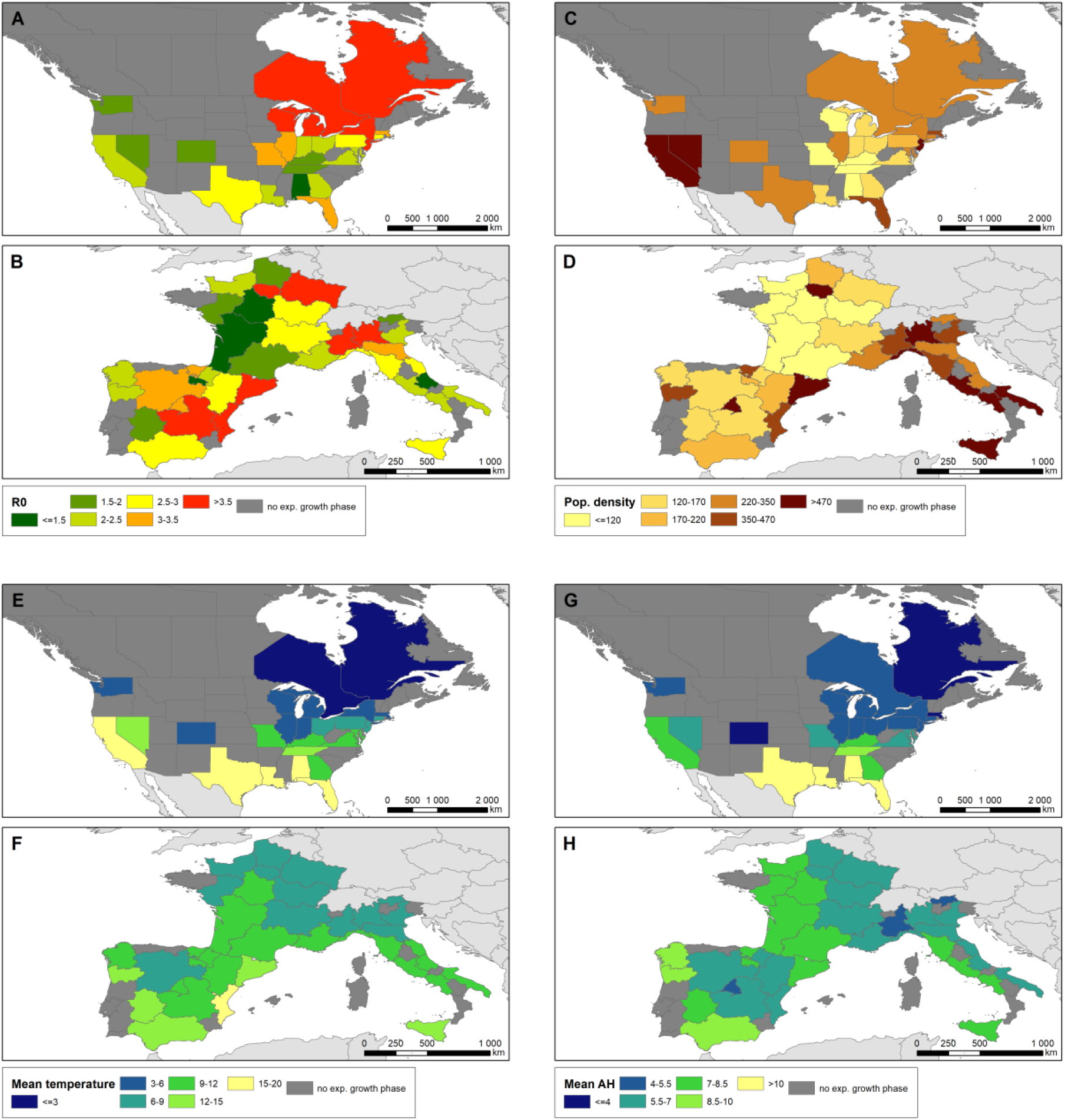
Map of regional values for R0 and selected covariates, panels are presented by continent. A,B: R0 ; C,D: population density (inhabitants per km2) ; D,E: mean temperature ; F,G: mean absolute humidity.

**Figure 4:**
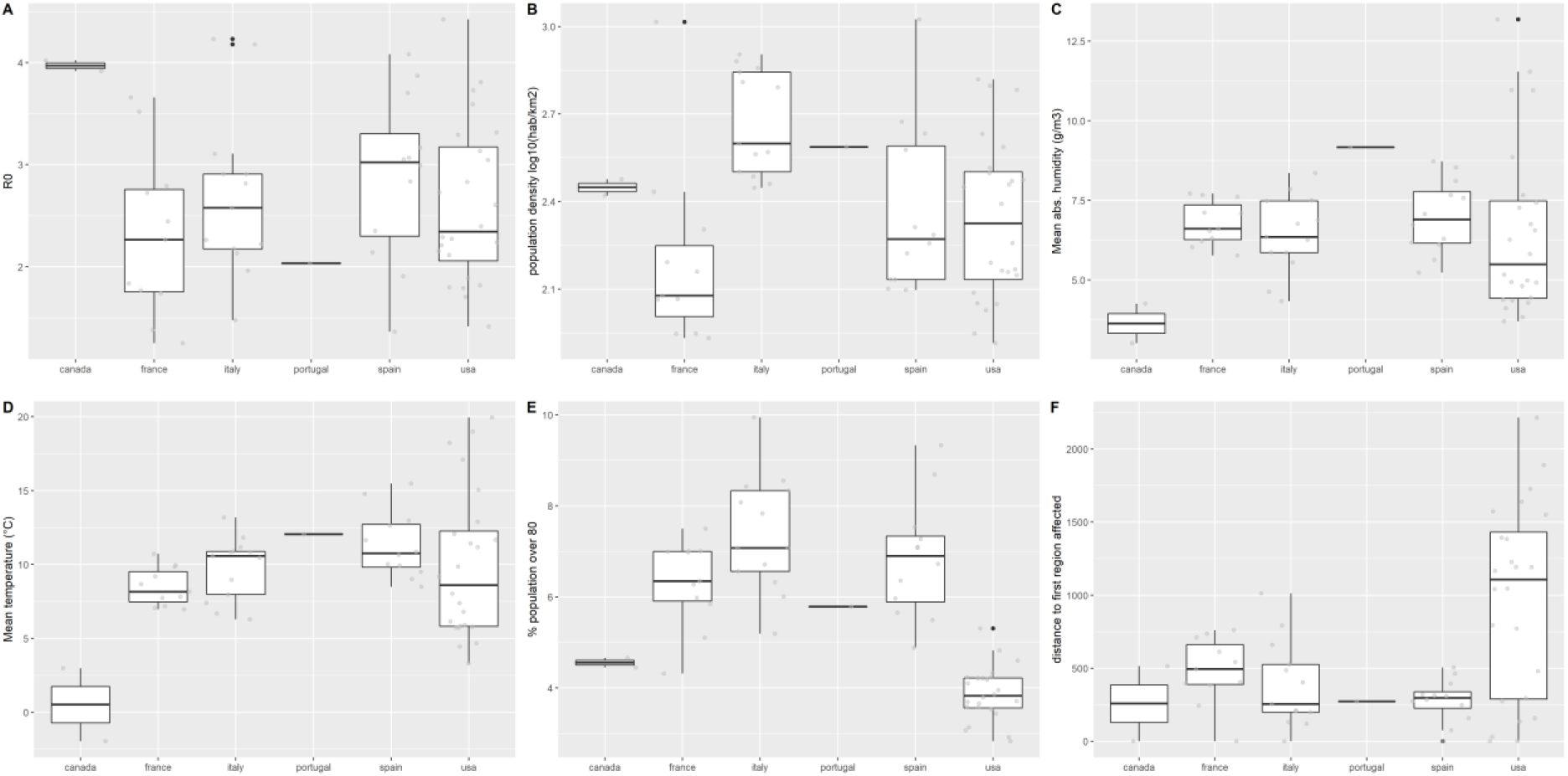
Distribution of R0 and selected covariates by country. A: R0; B: population density (log10 inhabitants/km2); C: Mean absolute humidity (g/m3); D: Mean temperature (°C); E: Population over 80 years old (%); F: distance to the first region affected (km). The box represents the interquartile range and the median; whiskers correspond to the minimum between highest value and 1.5 IQR; black dots to outliers. All observations are ploted in light grey.

Covariates were heterogeneous between countries (Figure 3 and 4). Population density ranged between 82 and 1056 inhabitants/km^2^ (median=271, Figure 3C and D) and was overall higher in Italy (Figure 4B). Mean absolute humidity during the transmission period was 4.98g/m3 in median (IQR=4.29-5.99, range=2.26-11.32, Figure 3E and F) corresponding to a median dew point of 3.7°C (IQR=0.9-6.3, range=-7.7 – +15.3). Mean temperature was 9.8°C in median (IQR=7.1-11.5, range=-2.0-19.9). Distance to the first region with 10 cumulative deaths was 406km in median (IQR=228-794) and much larger in the USA compared to European countries (Figure 4F).

### Factors associated with R0

Each variable was included in a univariate model assuming linear (Figure 5, Table S2) or non-linear (spline smoothing, Table S3) relationships with R0. We identified significant relationships between R0 value and temperature, absolute humidity or dew point, average daily rainfall, but not with mean wind speed over the transmission period.

**Figure 5:**
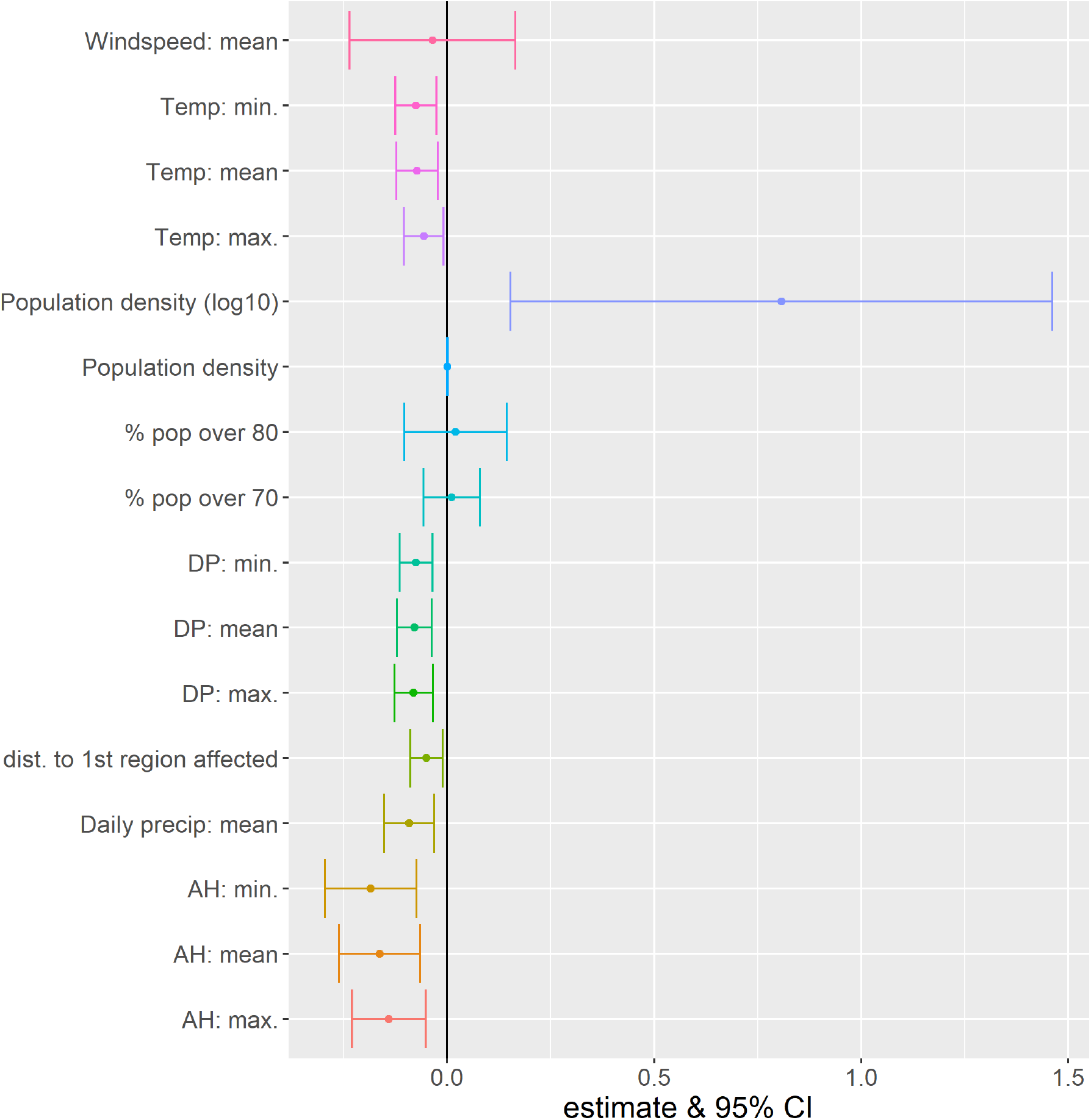
Change in R0 on an additive scale estimated from the univariable model assuming a linear relationship between R0 and the different variables.

Multivariable models were constructed according to the DAG. After adjustment for distance to the nearest affected region, percentage of population over 80 and population density, there was a consistent relationship between increasing temperature, AH or dew point temperature, and decreasing R0. No relationship was found with average daily rainfall.

Mean temperature led to the model with the largest deviance explained compared to models including minimum or maximum temperature, or any AH or dew point temperature (Table 1). In this model, a 10-fold (+1 log10 unit) increase in population density was associated with a +0.67 R0 unit increase (95%CI=0.05-1.28). The proportion of population aged 80 years and older was not significantly associated with R0. The relationship between mean temperature and R0 was not linear. A strong, nearly linear drop of approximately 1.0 R0 unit was observed between 2.5 and 12°C, and a plateau beyond 12°C (Figure 6 and 7). The residuals from this model did not exhibit significant spatial autocorrelation (Moran’s I=0.054, p=0.215).

**Table 1:** Multivariable model results for the relationship between R0 and weather parameters, obtained with the hierarchical generalized additive model. Weather parameters are temperature, absolute humidity, and dew point temperature, adjusted for distance to the first region affected, population density, and elderly population. Non-linear effects are presented in Figure 6. Models assuming linear effects for weather covariates are presented in Table S5.

| Model | Variable | Estimate | 95%CI | p-value |
| --- | --- | --- | --- | --- |
| **Model 1** | Intercept | 0.78 | [-0.88 - 2.45] | 0.36152 |
|  | Population density (log10) | 0.67 | [0.07 - 1.26] | 0.03218 |
|  | % population over 80 | 0.05 | [-0.08 - 0.18] | 0.44470 |
|  | Distance to first region affected in the country/coast | spline |  | 0.08007 |
|  | **Mean temperature** | spline |  | 0.00655 |
|  | Dev. explained: 41.5% |  |  |  |
| **Model 2** | Intercept | 1.28 | [-0.36 - 2.92] | 0.13239 |
|  | Population density (log10) | 0.50 | [-0.11 - 1.11] | 0.11294 |
|  | % population over 80 | 0.03 | [-0.1 - 0.16] | 0.61862 |
|  | Distance to first region affected in the country/coast | spline |  |  |
|  | **Mean AH** | spline |  | 0.03401 |
|  | Dev. explained: 33.6% |  |  |  |
| **Model 3** | Intercept | 1.20 | [-0.47 - 2.88] | 0.16427 |
|  | Population density (log10) | 0.49 | [-0.12 - 1.1] | 0.12005 |
|  | % population over 80 | 0.05 | [-0.08 - 0.18] | 0.47092 |
|  | Distance to first region affected in the country/coast | spline |  | 0.09756 |
|  | **Mean Dew Point Temperature** | spline |  | 0.00494 |
|  | Dev. explained: 34.6% |  |  |  |

**Figure 6:**
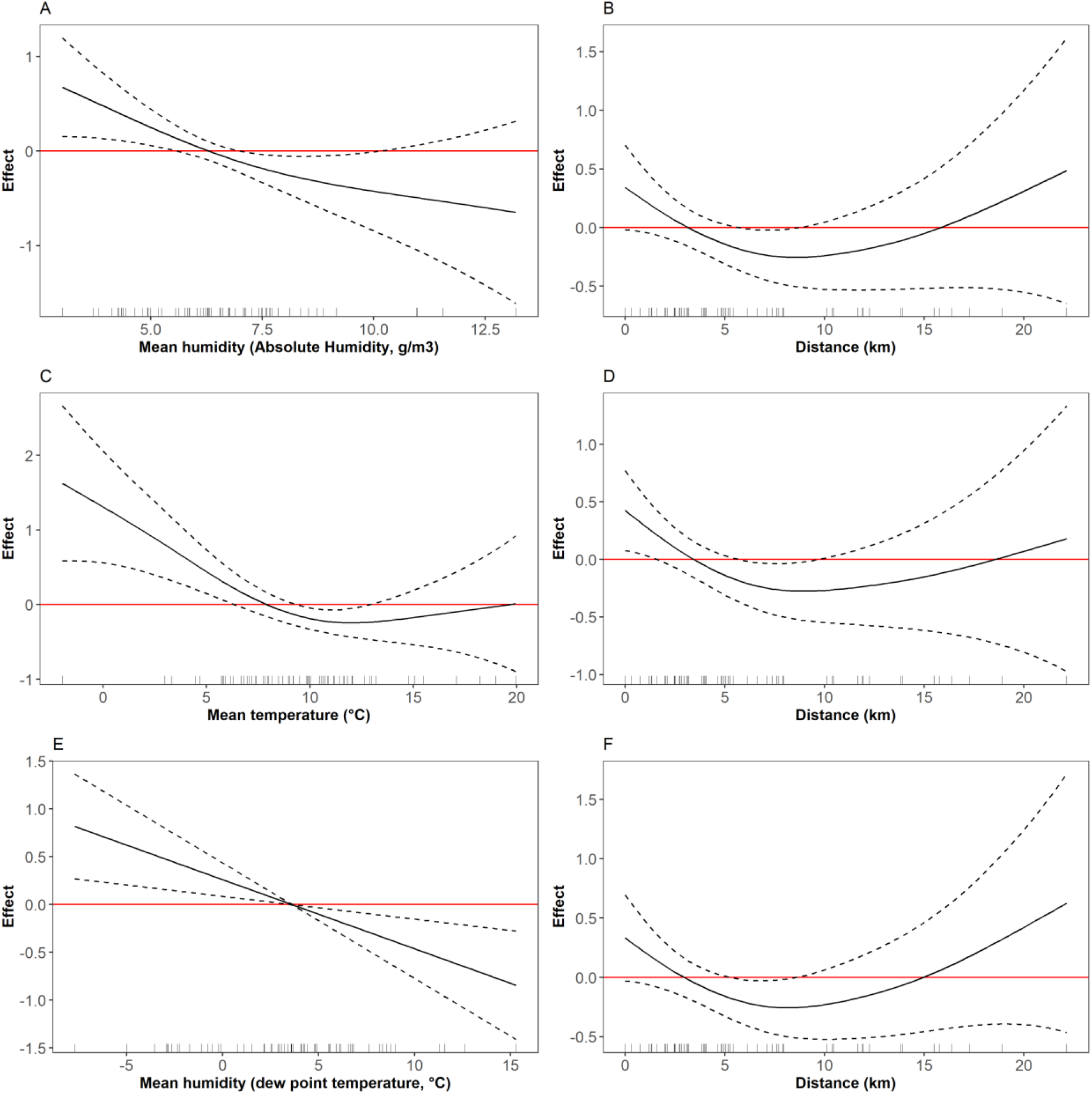
Non-linear effects in the multivariable model for weather parameters (see Table 1 for linear effects). A: Temperature, model 1. B: Distance to first region affected, model 1. C: absolute humidity, model 2. D: distance to first region affected, model 2. E: dew point temperature, model 3. F: distance to first region affected, model 3.

**Figure 7:**
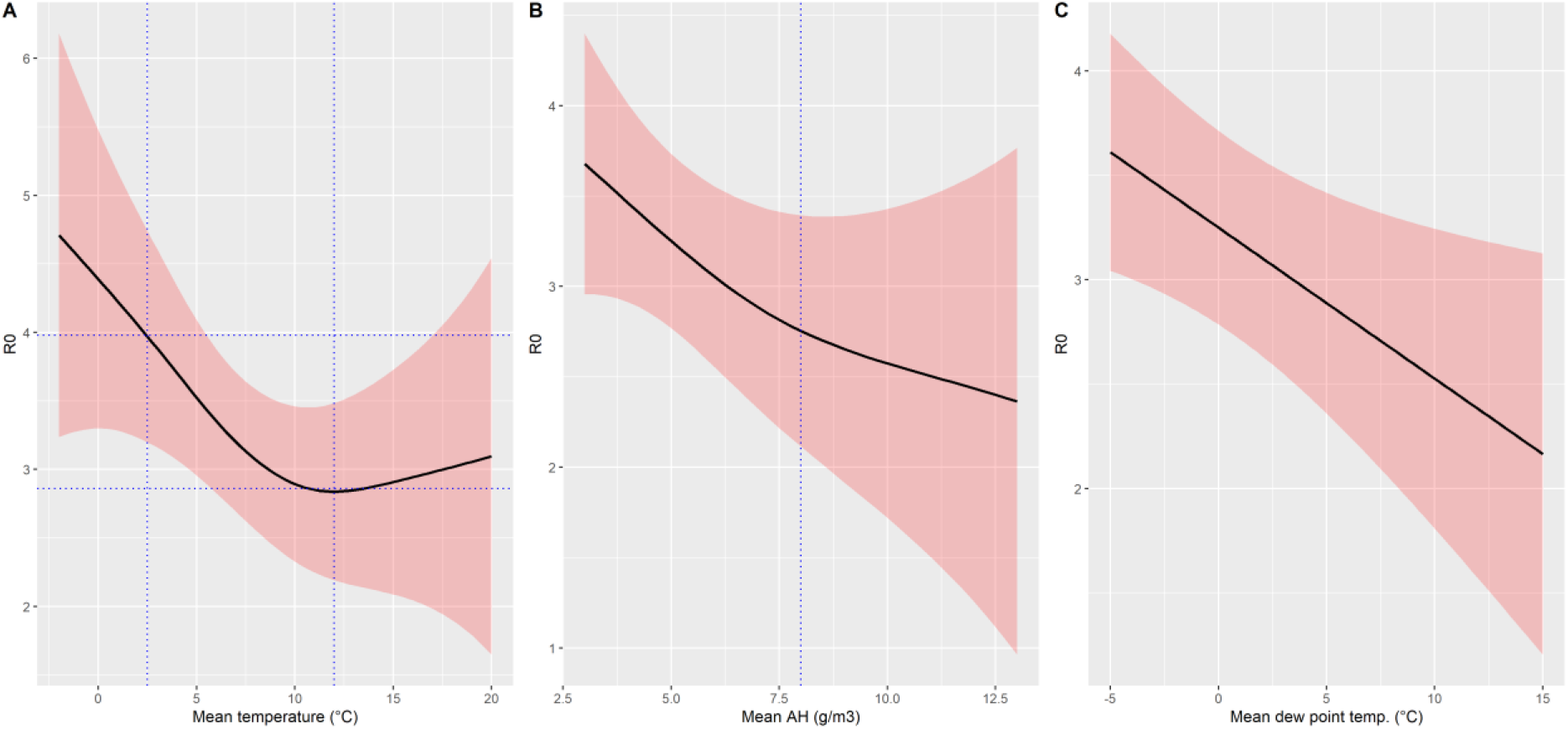
Summary of estimated effect of temperature (A), absolute humidity (B) or dew point temperature (C) on R0 assuming a region with average population density (248 persons/km2) and percentage of inhabitants >80 years (5.6%), and corresponding to the first region first affected (distance=0km).

Mean AH and mean dew point values exhibited similar profiles with increasing values associate with decreasing R0 values (Table 1, Figure 6). In spite of the narrower range of values, it seemed that the relationship between R0 and AH or dew point temperature did not reach a plateau (Figure 7). Assuming a linear relationship, a 1 g/m3 higher absolute humidity translated in a 0.15 unit lower regional R0, respectively a 1°C higher dew point temperature translated in a 0.08 unit lower R0 (Table S5).

### Sensitivity analyses

Using lagged weather summary values as linear univariate predictors, our statistical model found similar relationships between R0 and temperature variables, but 0-, 1- and 5-week lags led to the strongest effect for mean AH and mean DP (Figure S5). Using lagged weather summary values as non-linear predictors in the multivariate model, the shapes of the relationships between temperature, AH and DP remained similar, with a nearly linear drop reaching a plateau at values corresponding to milder winter weather/climate (Figure S6). Overall, correlations were strong between weather summary observations at the different lags (Figure S7). Correlations of lagged temperature and humidity observations was highest between -1, 0, 1 and 5-week lagged observations compared to other lags.

When setting the upper limit of the R0 calculation window to 18-day after date of lockdown, 10 additional regions were excluded and the analysis was conducted on 53 regions. After adjusting for population density, population over 80 and distance to the first region affected, the non-linear negative relationship between R0 and temperature remained unchanged, reaching a plateau around 10°C (spline p-value=0.0475). The shape of the relationship between AH, respectively DP, and R0 remained similar but did not reach statistical significance (p-value=0.19). Later AH or DP summary values, corresponding to 1 week later than the estimated transmission period (i.e. 2-week lag from R0 calculation period), restored the negative relationship with R0 (p=0.0661, respectively p=0.0667).

Fitting continent-specific splines for weather covariates, also resulted in low statistical power: only 37 regions remained in South Europe and 26 in North America. After adjusting for population density, population over 80 and distance to the first region affected, humidity variables (AH or dew point) retained a negative association with R0 (continent-specific spline p-values were 0.0915 (North America) and 0.0988 (South Europe) for AH, respectively 0.0506 and 0.1078 for dew point temperature). The relationship between R0 and mean temperature was markedly different between North America (negative association, p=0.005) and South Europe (p=0.17).

## Discussion

In this study, we analysed SARS-CoV-2 propagation parameter R0 during the first wave of the pandemic in 63 regions of 6 north western countries. We showed that R0 values were influenced by population density (+0.6 for a 10-fold increase in density), by proximity with the first epidemic focus of the country or coast for USA (−0.3 for a 10-fold increase in distance to the first region to record 10 COVID-19 deaths), and by weather or climate conditions. For regions with mean temperatures below 10°C during the transmission period, a linear association was observed with R0 values: a 1°C increase in temperature between regions was associated with a 0.16-unit decrease in R0. A 1 g/m^3^ increase in mean absolute humidity was associated with a 0.15-unit decrease in R0. Similar results were obtained with dew point temperatures, with a 1°C increase associated with a 0.08-unit decrease in R0 (Table S5). After adjusting for major confounders and spatial autocorrelation, our results indicate that weather conditions brought a significant contribution to drive the magnitude of the first wave, even if it was limited by an initial heterogeneous spread of the virus, which protected regions located furthest away from the first foci.

This study relied on a regional scale analysis and accounted for different dynamics within the same country. The overall epidemic wave at country level was actually the sum of diverse dynamics, as is obvious for large countries but also true for Spain, Italy or France, where regions were heterogeneously affected, as confirmed by serological studies [3, 17]. Likewise, weather or climate heterogeneity between regions of a given country was large. By analyzing distinct spatial units with heterogeneous population density and weather, we were able to assess the effect of parameters that may be otherwise confounded by country-level parameters such as response strategy, timing of the analysis period compared to the progression of the epidemic, but also age structure of the population.

The 6 countries were selected due to their homogeneous location in the northern hemisphere between 25 and 50 degrees of latitude and the low efficacy of their counter epidemic responses until a lockdown was decreed. We are therefore as close as possible of conditions allowing R0 estimation. This 4-parameter hierarchical generalized additive model provides an explanation of up to 45% of R0 variability across 63 affected regions of the northern hemisphere, thus providing useful insights on the drivers of the first wave, and allowing to estimate the contribution of population density and weather conditions for the next waves, when the effect of local introduction is no longer relevant.

We acknowledge several limits. The first one is the necessity to rely on death counts to estimate R0 during this wave due to the lack of information on actual infections from limited testing and the lack of consistency between countries for hospitalization counts. This assumption is similar to usual assumptions for R0 estimation based on diagnosed cases, since true number of infections remains unknown and detection occurs at variable delays after infection.

A second limit is the use of a single summary weather observation over the assumed transmission period, which may fail to express the dynamical aspects of weather. The choice of fixed, 1-week increments to study the effect variations in the weather summary window may be too coarse to capture effects for narrow exponential growth periods. The sensitivity analysis showed a slight increase in effect when considering weather summary values calculated 1 week earlier than the estimated transmission period, but the 18-day sensitivity analysis showed a stronger negative relationship between R0 and AH summary values corresponding to 1-week later than the estimated transmission period. The role of the weather conditions might be more important during the beginning of the exponential growth period, until a sufficiently large number of persons becomes infected and parameters such as population density become increasingly important. This study could also only evaluate the effect of a limited range of weather conditions on SARS-CoV-2 transmission, since the number of observations with a mean temperature below 2.5°C or above 15°C was low. This prevents from analyzing the effect of higher temperatures such as during autumn. This restriction was however necessary to achieve a minimal homogeneity of the studied regions in terms of their exposure and response to the pandemic. Finally, this analysis includes only 63 regions and may lack statistical power. The necessity to ensure that the epidemic growth of deaths was sufficient led to the exclusion of regions that may be affected, but not enough for daily deaths counts to reach 10 deaths/day.

Finally, this study showed an association between SARS-CoV-2 R0 and temperature/absolute humidity, but due to the strong correlation between absolute humidity and temperature in the seasonal conditions analysed here, it is difficult to determine which parameter is more important and they could not be analysed in combination [18]. The continent-specific analysis suggests that the relationship between absolute humidity and R0 was more stable than that of temperature, which was strong in the US but less so in South Europe. We could not conclude whether the relationship results from a direct role of weather on individual susceptibility to viral invasion (*e*.*g*. dry nasal mucosa from indoors heating and outdoors cold) or on viral persistence/survival, or from an indirect role of climate on human behaviors (*e*.*g*. regions with cold winter favor more indoors living conditions and lead to bigger infection opportunities).

## Conclusion

Our study shows an important dependency of SARS-CoV-2 transmission to weather/climate, with a 0.16-unit increase in R0 for a 1°C difference in mean regional temperature below 10°C, or a 0.15-unit increase for a -1g/m3 decrease in absolute humidity. Northern hemisphere countries experienced a second wave of SARS-CoV-2 infections during autumnal transition from summer to winter, while still actively maintaining control strategies. When planning to adjust the level of restrictions on social activities, public health strategies need to account for the increased transmissibility of SARS-CoV-2 when/where cold and dry winter conditions are prevalent.

## Material and methods

### Study design

In order to compare the drivers of the epidemic dynamics between regions accurately, we calculated the basic reproduction number (R0) of the virus for each region affected by the first wave of COVID-19 epidemic in six countries, using the dynamics of the daily death counts. These six countries were located in the western part of the northern hemisphere, approximately between 25 and 50° of latitude (Figure S1). These countries experienced winter conditions and underwent significant SARS-CoV-2 transmission in a context of low public health response at the start of the first wave of the epidemic.

This analysis was conducted at the first administrative subdivision country, here referred to as “region”. Regions corresponded to States in USA and Canada, autonomous communities in Spain, and regions in Italy (regioni), Portugal (região) and France (regions).

Confirmed case counts were insufficiently reliable due to overall lack of tests and different testing strategies between countries, between regions of the same country and between periods for the same region. Hospitalization counts were not available consistently at regional level across countries. Overall, COVID-19 deaths were preferred as they were less likely to have different definitions within the same region or country and to undergo significant changes over the study period. R0 is an indicator of the speed of progression of the outbreak, and was therefore less likely to be biased compared to indicators based on cumulative counts or cumulative incidence rates. Estimating R0 values based on deaths counts relies on the minimal assumption that the infection-fatality rate was constant over the study period (∼1 month) for a given region, which defined a proportional relationship between infections and deaths.

### Study period

We included COVID-19 death count data starting at the date when 10 cumulative deaths were reached in a given region, and ending 28 days after lockdown.

The lower boundary of 10 cumulative deaths was chosen to avoid early stochasticity and limitations in the available recordings of the first COVID-19 deaths at regional level.

The upper boundary of 28 days after lockdown was defined to avoid the influence of lockdown measures on the growth rate of death counts, since we aimed to estimate SARS-CoV-2 R0 prior to implementation of major interventions. At individual level, the median delay between infection and death was 18 days, with a large interquartile range (IQR) of 9 to 24 days [19–25]. At regional level, we assumed that reported deaths corresponded to transmission events which had occurred in median 3 weeks earlier, and defined a 28-day boundary to cover the upper limit of the IQR.

### Data

#### Deaths from COVID-19

Regional level data on COVID-19 deaths were retrieved from data shared by national health ministries and/or public health agencies of Spain, Italy, Portugal, France, United States of America, and Canada (Table S1). Deaths were reported as daily new death counts or as a cumulative number.

#### Population and geographical data

Population structure by one- or five-year age groups, sex and region was retrieved from open access data shared by national institutes or administrations responsible for national statistics or demography (Table S1). The percentage of the region population aged ≥70 or ≥80 years was calculated in order to adjust for differences in way of life (e.g. rural regions have older population than metropolitan areas in Europe).

Region shapefiles by country were obtained from national geographical authorities, open source datasets or as provided in the coronavirus open data packages proposed by several national health agencies (Table S1). The region surface was either obtained from the geographical layer (land surface area), or calculated from the polygon extent. We estimated the percentage of each region surface with ≥5 inhabitants per km^2^ based on WorldPop 2020 raster dataset [26]. Population density was estimated as the population in the region divided by the surface after excluding areas <5 inhabitants/km^2^ in order to limit the underestimation of population density when high heterogeneity existed between urban centers and sparsely populated territories within the same region (deserts, mountains, polar regions).

#### Weather/climate

Weather station reports since 1 January 2020 were obtained from US National Oceanic and Atmospheric Administration (NOAA) through the R-package {worldmet}. We extracted hourly temperature, relative humidity (RH), dew point (DP), precipitation and windspeed observations for each station. Hourly absolute humidity (AH, in g/m^3^) was calculated from RH and temperature based on the Clausius-Clapeyron formula [27]. Daily minimum, maximum and mean values were calculated for temperature, AH, RH, dew point, as well as cumulative sum of precipitation and mean wind speed. Days with ≥ 5 missing hourly record (no observation of any parameter) were excluded.

Each region was attributed stations based on geographic location. All available observations in weather stations of the region contributed to the regional daily average. Weather stations can be located in mountains or inhabited locations where they record weather conditions that differ strongly from actual populated areas of the same region. To avoid this bias, observations were assigned population-based weights: we estimated the population located within 10 km of each weather station using WorldPop 2020 data; and for each day and each region, we calculated the total population within 10km of any station reporting data for that day. Daily station observations were weighted in proportion of the population around each station relative to the total population. Stations located within 10-km of each other were included in a single buffer and each station was assigned equal weight within the buffer.

For each weather parameter, the regional summary value was calculated over the transmission period, during which infections were assumed to have occurred. We assumed that the transmission period had the same duration as the R0 calculation period, and occurred 3 weeks earlier according to the delay between infection and death (Figure S8).

Using this approach, the weather parameters averaged over the assumed transmission period and over each region included: minimum, mean and maximum average values for temperature, relative humidity, absolute humidity, dew point, average cumulative rainfall per day and average wind speed.

### Date of lockdown definition

Lockdown date was defined using Google mobility data as the date when a decrease >25% in workplace localization was reported and sustained over 3 days in the region [28]. All references to a « date of lockdown » hereafter refer to this definition. This definition matched national lockdowns in European countries. This simplification was necessary due to the heterogeneity in social distancing measures taken at regional (state) level in the USA and Canada. The objective was to exclude periods where transmission would start to slow down due to these measures.

### Distance to first region with 10 cumulative deaths

For each of the four European countries considered, we identified the first region with 10 cumulative deaths was defined. In Portugal, two regions reached ≥ 10 deaths on the same day, and the region with the highest count (14 versus 12) was selected. Within each country, the euclidean distance in kilometers between the main city in each region and the main city in the first region above 10 cumulative deaths was calculated to reflect spatial autocorrelation due to proximity in the spread of the epidemic. In the USA and Canada, the distance to the first region above 10 cumulative deaths was calculated separately for East Coast and West Coast, using the limit between Central and Mountain time zones. This was necessary due to the early start of the epidemic in the state of Washington, which reached 10 deaths by 2 March 2020, 16 days before the next state (New York on 18 March).

### Statistical methods

Statistical analyses were performed using R version 4.0. Maps were produced using ArcGIS 10.7.1.

### R0 estimation

Daily death counts were smoothed using a 5-day moving average filter to account for irregularities in data transmission and publication.

For each region, the exponential growth period was estimated using a log(deaths)=f(time) representation and r (the exponential growth rate) was extracted as the coefficient of a Poisson regression. R0 was calculated for each region using the generation time method assuming a gamma distribution with parameters 7 and 5.2 for SARS-CoV-2 generation time [29, 30]. In order to improve the adjustment of the regression, the start and end dates of the calculation period were allowed to shift by up to 2 days (+/-1 day or +1/+2 days for start date if the calculation period began at the date of 10 cumulative deaths) using the built-in function sensitivity.analysis() of the package {R0} [31].

### Exclusion criteria

Regions where the smoothed daily death count stayed <5 deaths/day during the study period were not included. Region where the smoothed daily death count stayed <10 deaths/day during the linear growth phase were excluded. This limited the study to regions having displayed a clear exponential growth phase.

One Italian region was excluded because there was no weather station data available except for 2 stations >2000m altitude.

### Analysis of the relationship between R0 and weather parameters

A directed acyclic graph (DAG) was constructed using Dagitty v3.0 web-based application (http://www.dagitty.net/dags.html) in order to visualise the relationships between R0 and the explanatory covariates (Figure S2). Dependence and independence assumptions were verified using Spearman correlation coefficient.

Relative humidity values are temperature-dependent and absolute humidity or dew point temperatures are also strongly correlated to temperature [18]. The different weather covariates were observed during the estimated transmission period, which corresponded to the R0 calculation period lagged by 3 weeks to account for delay between infection and death. Weather covariates were included separately in the models. A generalized additive mixed model (gamm) regression was used to evaluate the effects of climate, population and other determinants on the value of R0, using a Gaussian distribution and the identity link function (package {mgcv}). A country-level random effect was included to account for within-country correlations. Canada presented with only 2 regions and was grouped with the USA for random effect, while the single region in Portugal was grouped with Spain. Univariate analyses were conducted assuming linear and non-linear effects, using B-splines to model non-linear effect of covariates. Models were compared using the percentage of deviance explained and Akaike’s Information Criterion.

We verified presence/absence of spatial autocorrelation in R0 values and in the final model residuals by calculating Moran’s I.

### Sensitivity analyses

First, we assessed the importance of the 3-week delay for weather variables (corresponding to the delay between transmission period and R0 calculation period based on death count exponential growth). For this, we tested a variety of lags from -1 to 5 weeks from that estimated transmission period (*i*.*e*. 2 to 8 weeks from the R0 calculation period). The longer lags were likely irrelevant for actual transmission but aimed at identifying climate trends rather than weather influence. We included them as linear explanatory variables in the univariable hierarchical model or as non-linear explanatory variables in the multivariable hierarchical generalised additive model.

Second, we assessed the effect of the 28-day window to define the exponential growth period by using a narrower window ending 18 days after date of lockdown. We recalculated R0 for regions where the linear growth period retained for the main analysis extended beyond the 18-day limit, and followed the same plan as the main analysis.

Third, we assessed possible continent specific effects by fitting continent-specific splines for weather variables in the final multivariate models.

### Data availability

All data used in this study was obtained from publicly available data sources, listed in Table S1. The only exception corresponded to early death counts at regional level in France which were obtained directly from the French Public Health Agency but have been publicly released since. The data table of regional-level values (R0 estimates, weather summary, population density etc) used for the hierarchical generalized additive model analysis of the relationship between regional level R0 and weather covariates is provided in a supplementary csv file.

### Code availability

No custom code was used for this study beyond usual application of standard functions of softwares or R packages.

## Supporting information

Appendix file containing supplementary materials

## Data Availability

The analysis was performed using openly accessible data. The links are provided in the supplementary material.

## Authors’ contribution

JL, JG, SC and AF designed the study. JL conducted the analysis with methodological contributions of JP, EL, and LL. JL JP EL LL AF SC JG contributed to interpreting the results and editing the manuscript.

## Acknowledgements

The authors would like to thank Laurax Simgiane Ferbliegas from Hab’ lab Marseille and L. Jae for help and support in retrieving multi-country regional data.

