## Appendix file containing supplementary materials for "Colder and drier winter conditions are associated with greater SARS-CoV-2 transmission: a regional study of the first epidemic wave in north-west hemisphere countries"

Version 1.1, 27 January 2021

Jordi Landier[1*], Juliette Paireau[2,3], Stanislas Rebaudet[1,4], Eva Legendre[1], Laurent Lehot[1], Arnaud Fontanet[5,6], Simon Cauchemez[2], Jean Gaudart[7]

[1] IRD, Aix Marseille Univ, INSERM, SESSTIM, Marseille, France.

[2] Mathematical Modelling of Infectious Diseases Unit, Institut Pasteur, UMR2000, CNRS, Paris, France.

[3] Santé publique France, French National Public Health Agency, Saint Maurice, France

[4] Hôpital Européen Marseille, France.

[5] Emerging Infectious Diseases Unit, Institut Pasteur, Paris, France.

[6] PACRI Unit, Conservatoire National des Arts et Métiers, Paris, France.

[7] Aix Marseille Univ, APHM, INSERM, IRD, SESSTIM, Hop Timone, BioSTIC, Marseille, France.

**Systematic review of available evidence**

Systematic reviews addressed the issue until 15 May 2020 (Briz-Redon A, et al, Prog Phy Geogr 2020 & Mecenas P et al, PLoS One, 2020).

Between 15 May 2020 and 15 December 2020, we searched PubMed database using the request: “(((COVID-19 OR SARS-COV-2)) AND (("2020/05/16"[Date - Publication]: "2020-12-15"[Date - Publication]))) AND (humidity [MeSH Terms] OR temperature [MeSH Terms] OR weather [MeSH Terms]OR climate[MeSH Terms])”. Of 464 references, we identified 82 research articles relative to the relationship between weather parameters and SARS-CoV-2 transmission. Of these, 67 (82%) analysed case, death or hospitalization counts, of which 11 presented the analysis of a single time series and 56 the analysis of cumulative or longitudinal counts in multiple locations ranging from regions to countries. Of 67, 52 (78%) presented only univariate analyses of weather variables, or the analysis of multiple correlated weather variables without evaluation of potential confounders. Only 15 (18%) studies analysed the growth rate or the reproduction number of SARS-CoV-2, of which only 4 studies did a multivariate analysis.

In spite of their large numbers, univariate studies and studies based on counts suffered from multiple biases and, according to systematic reviews, provided only low-grade evidence of a negative relationship between SARS-CoV-2 and temperature.

**Table S1: Data sources**

| Country | Data type | source | source type | date accessed (or last update) | link |
|---|---|---|---|---|---|
| USA | deaths | The Covid Tracking Project | non governmental | 29/09/2020 | https://covidtracking.com/api/v1/states/daily.csv |
| USA | population | US Census Bureau | governmental | 22/04/2020 | https://www.census.gov/data/tables/time-series/demo/popest/2010s-state-detail.html |
| USA | shapefile | US Census Bureau | governmental | 23/04/2020 | https://www2.census.gov/geo/tiger/GENZ2018/shp/cb_2018_us_state_5m.zip |
| Canada | deaths | Public Health Infobase, Govt of Canada | governmental | 15/05/2020 | https://health-infobase.canada.ca/covid-19/ |
| Canada | population | Statistics Canada | governmental | 29/04/2020 | https://www12.statcan.gc.ca/datasets/index-eng.cfm?Temporal=2016 |
| Canada | shapefile | IGISMAP | non governmental | 30/04/2020 | https://map.igismap.com/share-map/export-layer/Canada_AL263/45fbc6d3e05ebd93369ce542e8f2322d |
| Italy | deaths | Protezione civile (Govt of Italy) | governmental | 24/09/2020 | https://raw.githubusercontent.com/pcm-dpc/COVID-19/master/dati-regioni/dpc-covid19-ita-regioni.csv |
| Italy | population | I.Stat | governmental | 23/03/2020 | http://dati.istat.it/Index.aspx?DataSetCode=DCIS_POPRES1 |
| Italy | shapefile | I.Stat | governmental | 23/04/2020 | https://www.istat.it/storage/cartografia/confini_amministrativi/generalizzati/Limiti01012020_g.zip |
| Spain | deaths | Centro Nacional de Epidemiologia | governmental | 25/05/2020 | https://covid19.isciii.es/ |
| Spain | population | Instituto Nacional d'Estadistica | governmental | 20/04/2020 | https://www.ine.es/jaxiT3/Datos.htm?t=31304#!tabs-mapa |
| Spain | shapefile | GADM | non governmental | 20/04/2020 | https://gadm.org/maps/ESP_1.html |
| Portugal | deaths | Data Science for Social Good Portugal | non governmental | 21/10/2020 | https://raw.githubusercontent.com/dssg-pt/covid19pt-data/master/data.csv |
| Portugal | population | Instituto Nacional d'Estadistica | governmental | 21/04/2020 | https://www.ine.pt/xportal/xmain?xpid=INE&xpgid=ine_indicadores&contecto=pi&indOcorrCod=0008273&selTab=tab0 |
| Portugal | shapefile | Data Science for Social Good Portugal | non governmental | 02/06/2020 | https://github.com/dssg-pt/covid19pt-data/blob/master/extra/mapas/portugal/portugal.shp |
| France | deaths | Santé publique France/SI-VIC | governmental | 24/04/2020 | Not available in open access |
| France | population | INSEE | governmental | 07/05/2020 | https://www.insee.fr/fr/statistiques/1893198 |
| France | shapefile | data.gouv.fr | governmental | 02/06/2020 | https://www.data.gouv.fr/fr/datasets/r/aacf9338-8944-4513-a7b9-4cd7c2db2fa9 |

**Figure S1: Study regions in six countries of the North West hemisphere and selection criteria**

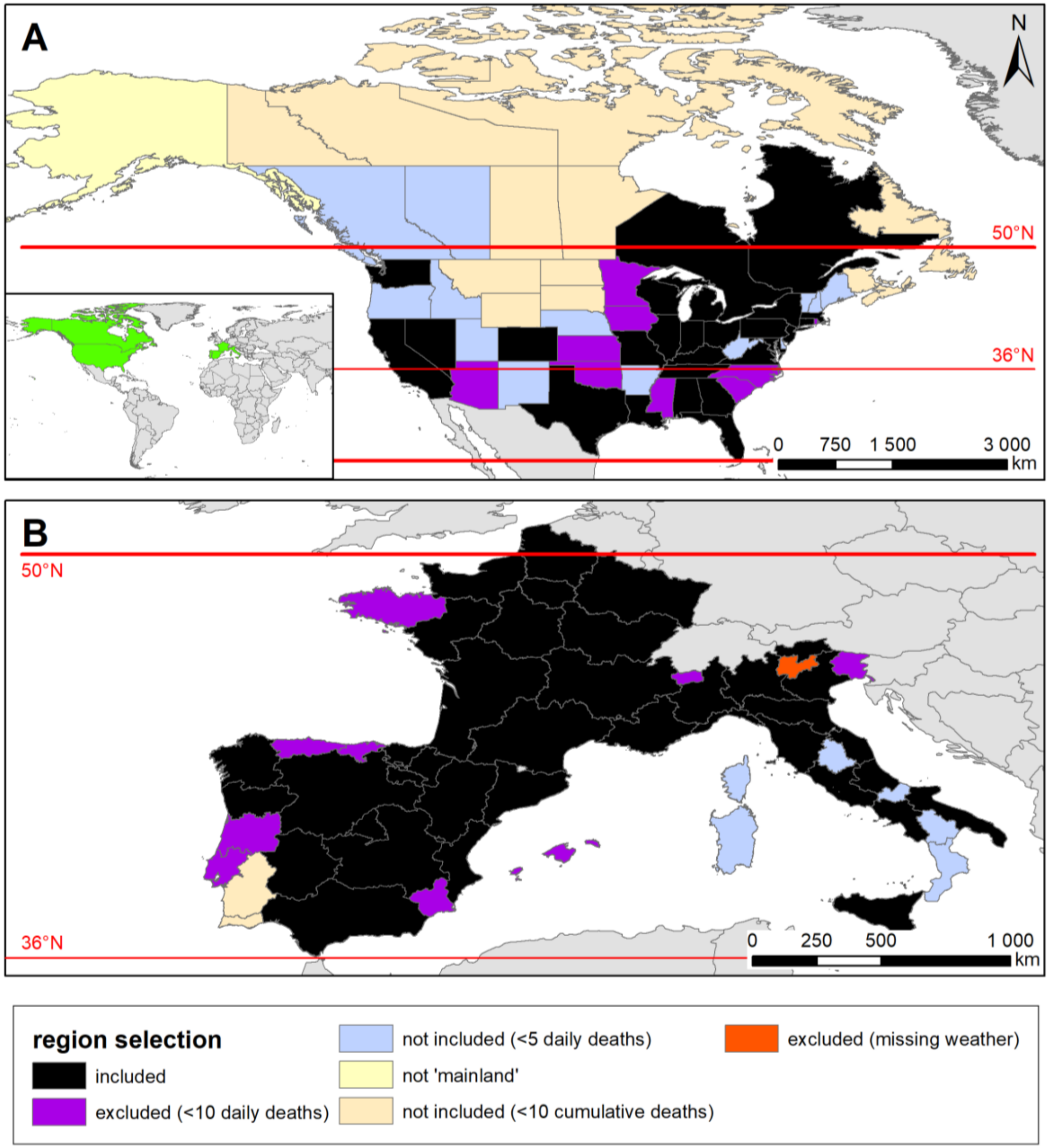

**Figure S2: Directed acyclic graph for the relationship between weather/climate parameters and SARS-COV2 transmission as measured by R0**

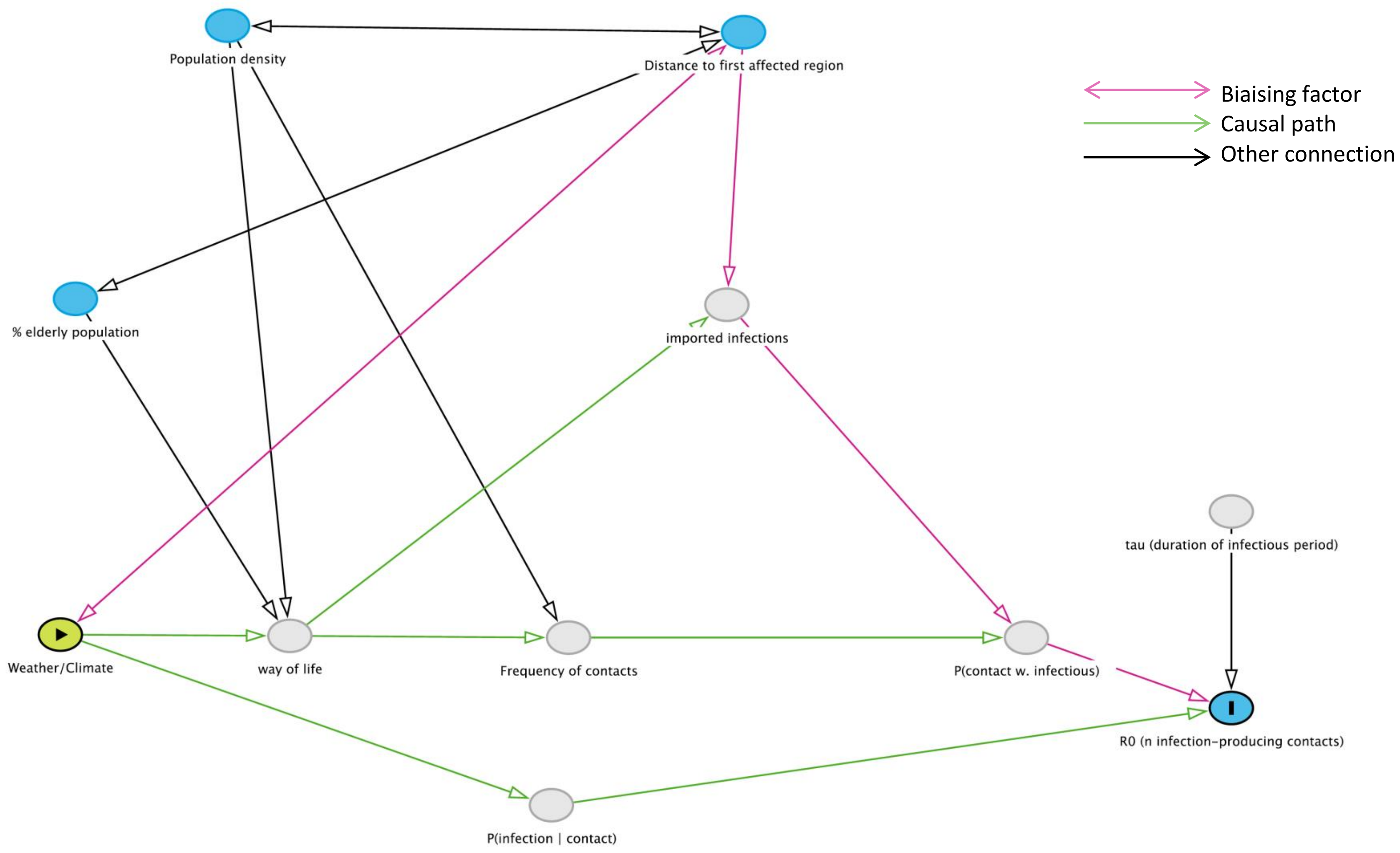

**Figure S3: delay to peak in daily deaths according to the reduction of population mobility measured in transit stations (A) or in workplaces (B). The death count at the peak (maximal death count) is figured by the dot size (0, 100, 400).**

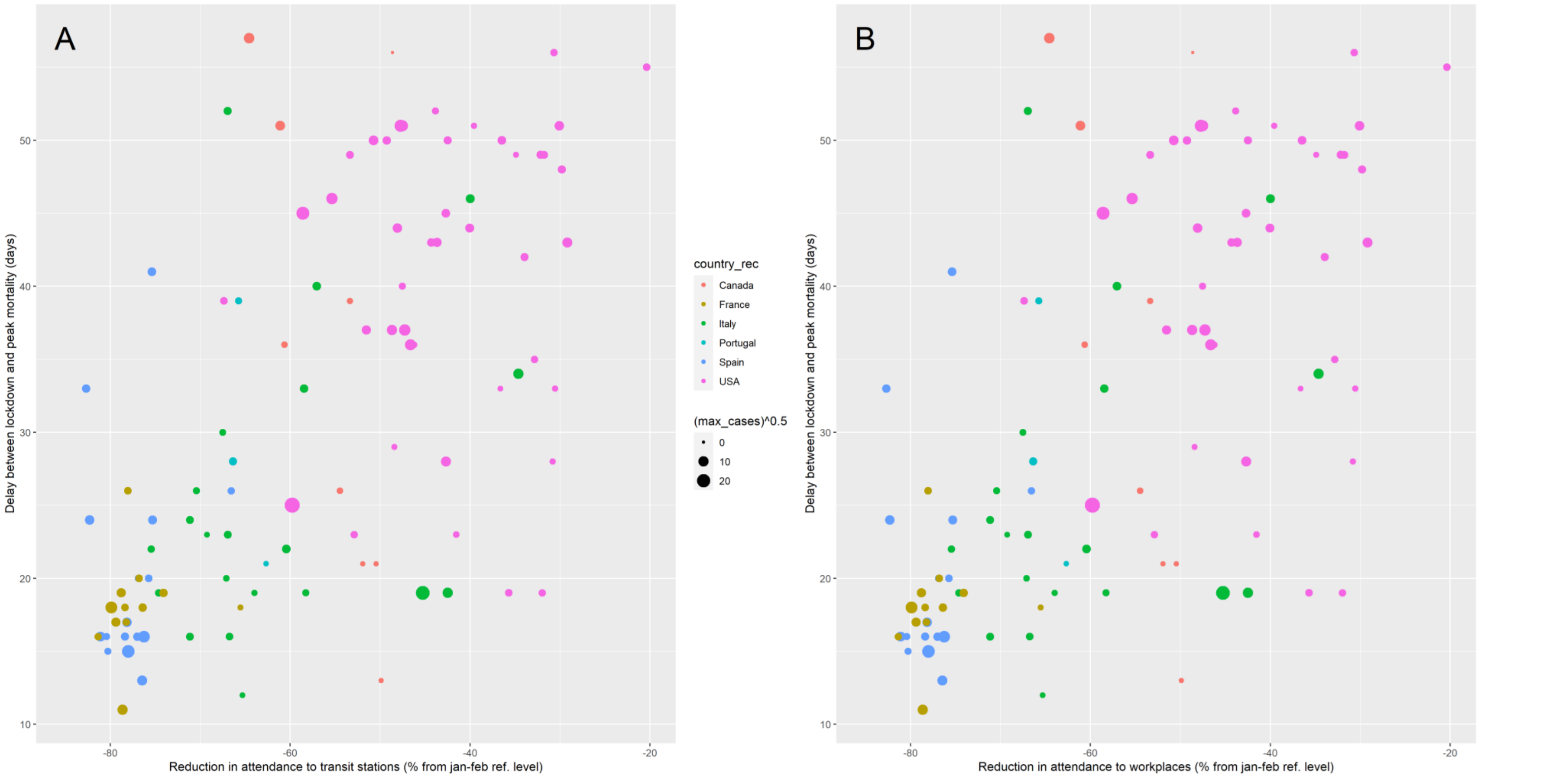

The correlation between the delay to reaching the peak in daily death counts and the reduction of population mobility was strong, with respective Spearman correlation coefficient values of 0.636 (p<10^-5) for transit stations and 0.631 (p<10^-5) for workplaces (p<10^-5).

**Figure S4: R0 calculation periods by country**

Vertical green lines indicate the window considered for R0 calculation, starting at the date of 10 cumulative deaths and ending 28 days after lockdown. Vertical blue line indicate the end of the window considered for R0 calculation in the sensitivity analysis, 18 days after lockdown. The portion of the epidemic curve defined as displaying linear trend on the log-scale and used to extract the growth rate and perform the R0 calculation is highlighted in red, and the corresponding linear regression is displayed in orange. (See Results section for median and IQR values for calculation periods).

#### France

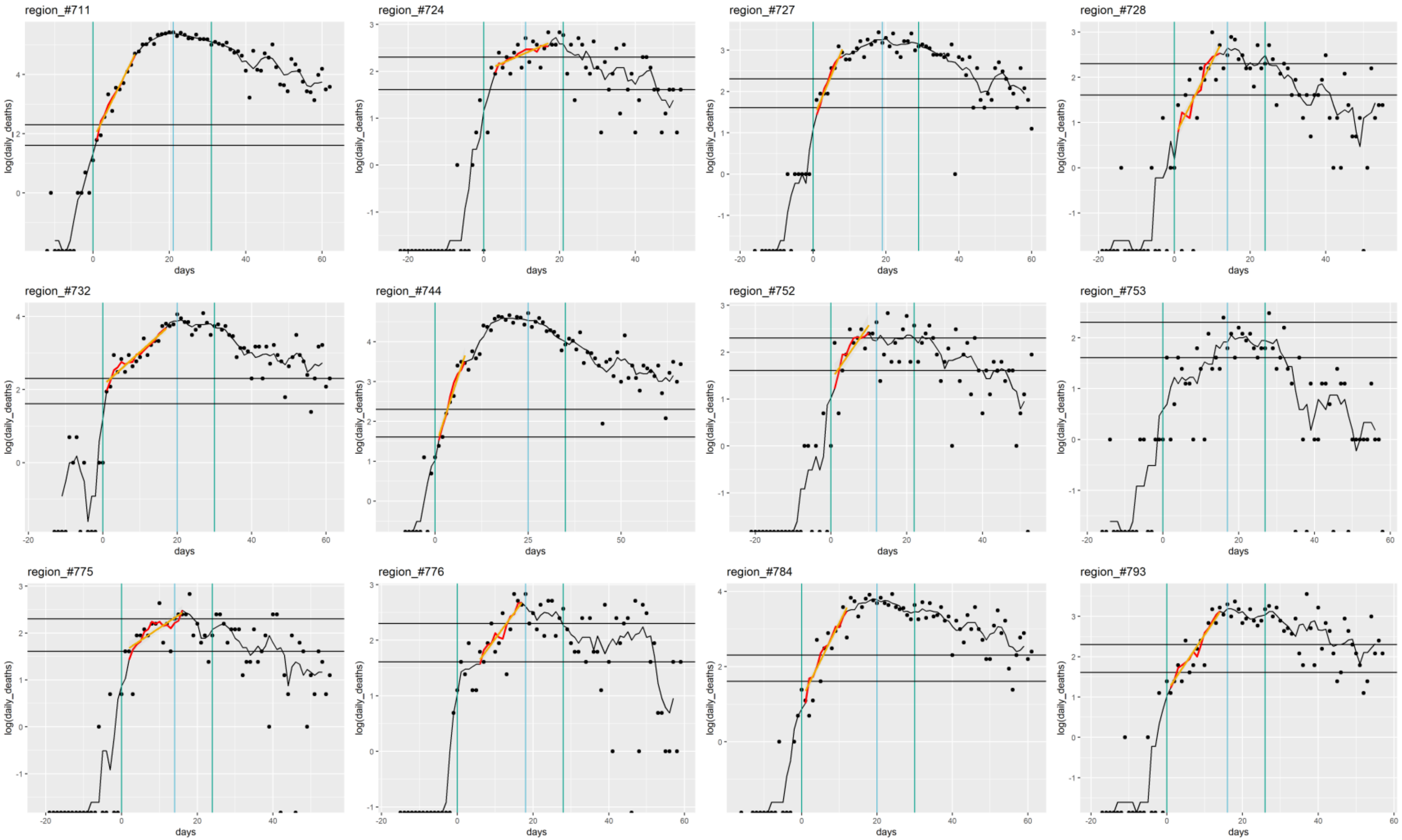
region_#711
region_#724
region_#727
region_#728
region_#732
region_#744
region_#752
region_#753
region_#775
region_#776
region_#784
region_#793
log(daily_deaths)
days

#### Italy

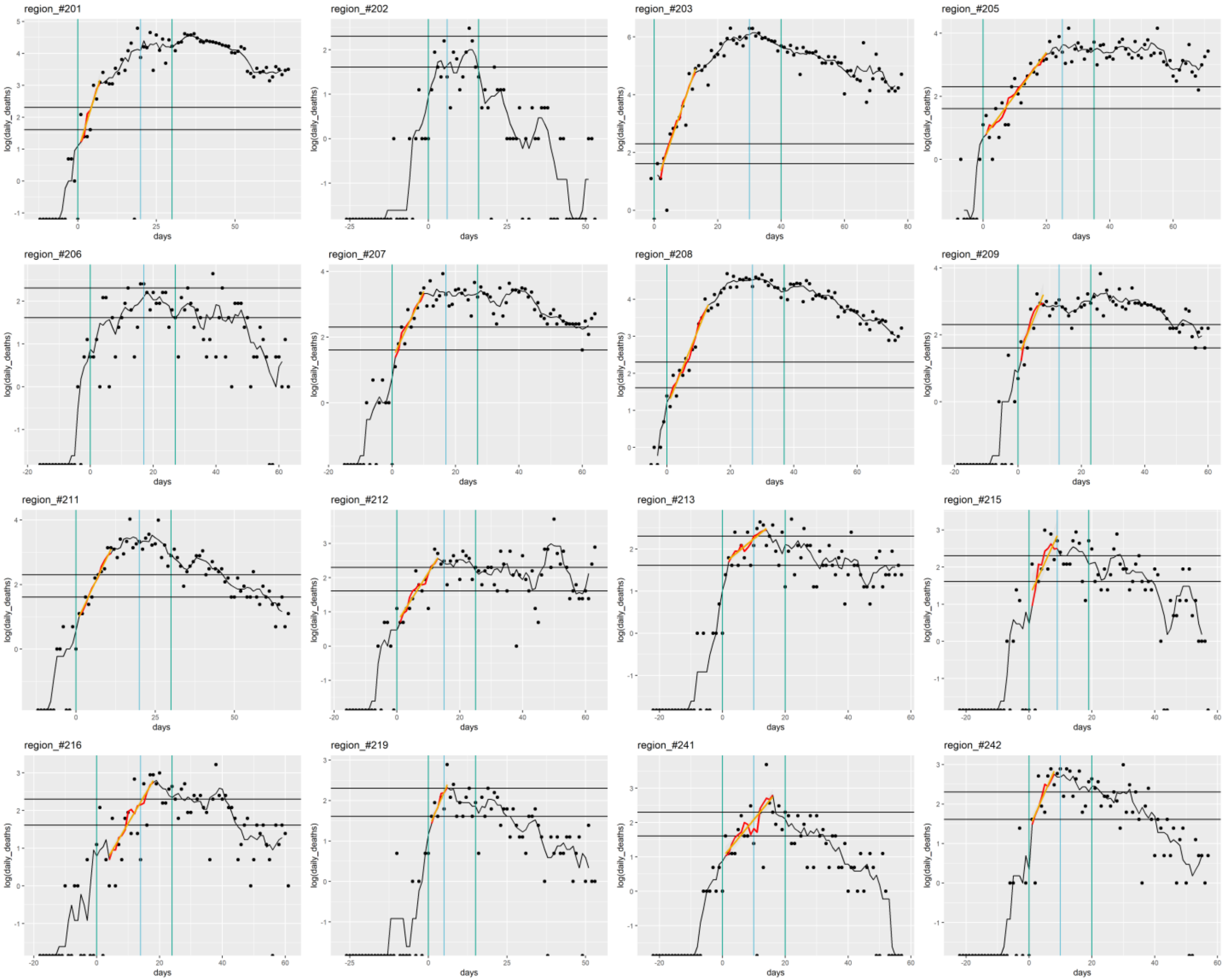
region_#201
region_#202
region_#203
region_#205
region_#206
region_#207
region_#208
region_#209
region_#211
region_#212
region_#213
region_#215
region_#216
region_#219
region_#241
region_#242
log(daily_deaths)
days

#### Spain

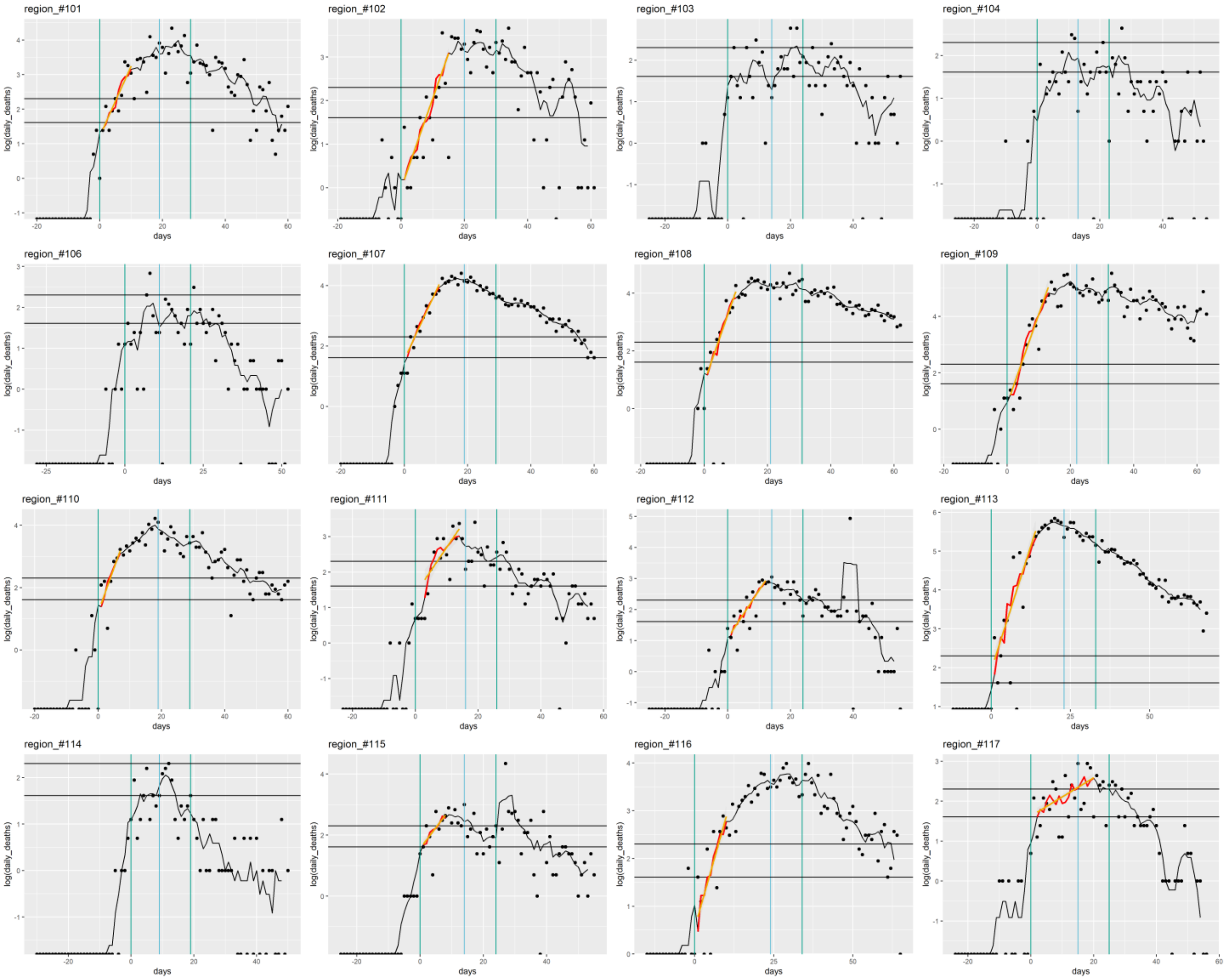
region_#101
region_#102
region_#103
region_#104
region_#106
region_#107
region_#108
region_#109
region_#110
region_#111
region_#112
region_#113
region_#114
region_#115
region_#116
region_#117
log(daily_deaths)
days

### USA

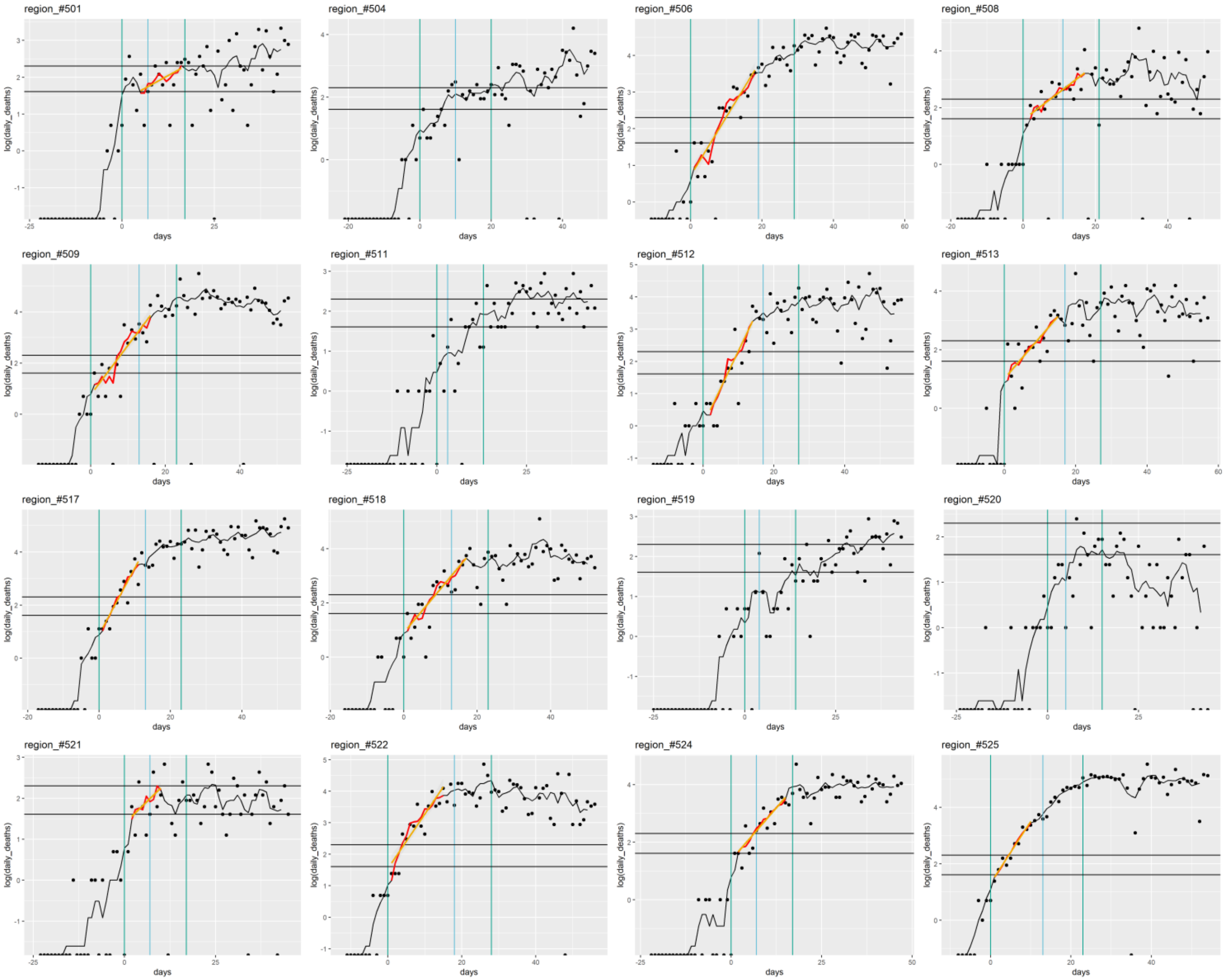
region_#501
region_#504
region_#506
region_#508
region_#509
region_#511
region_#512
region_#513
region_#517
region_#518
region_#519
region_#520
region_#521
region_#522
region_#524
region_#525
log(daily_deaths)
days

#### USA (continued)

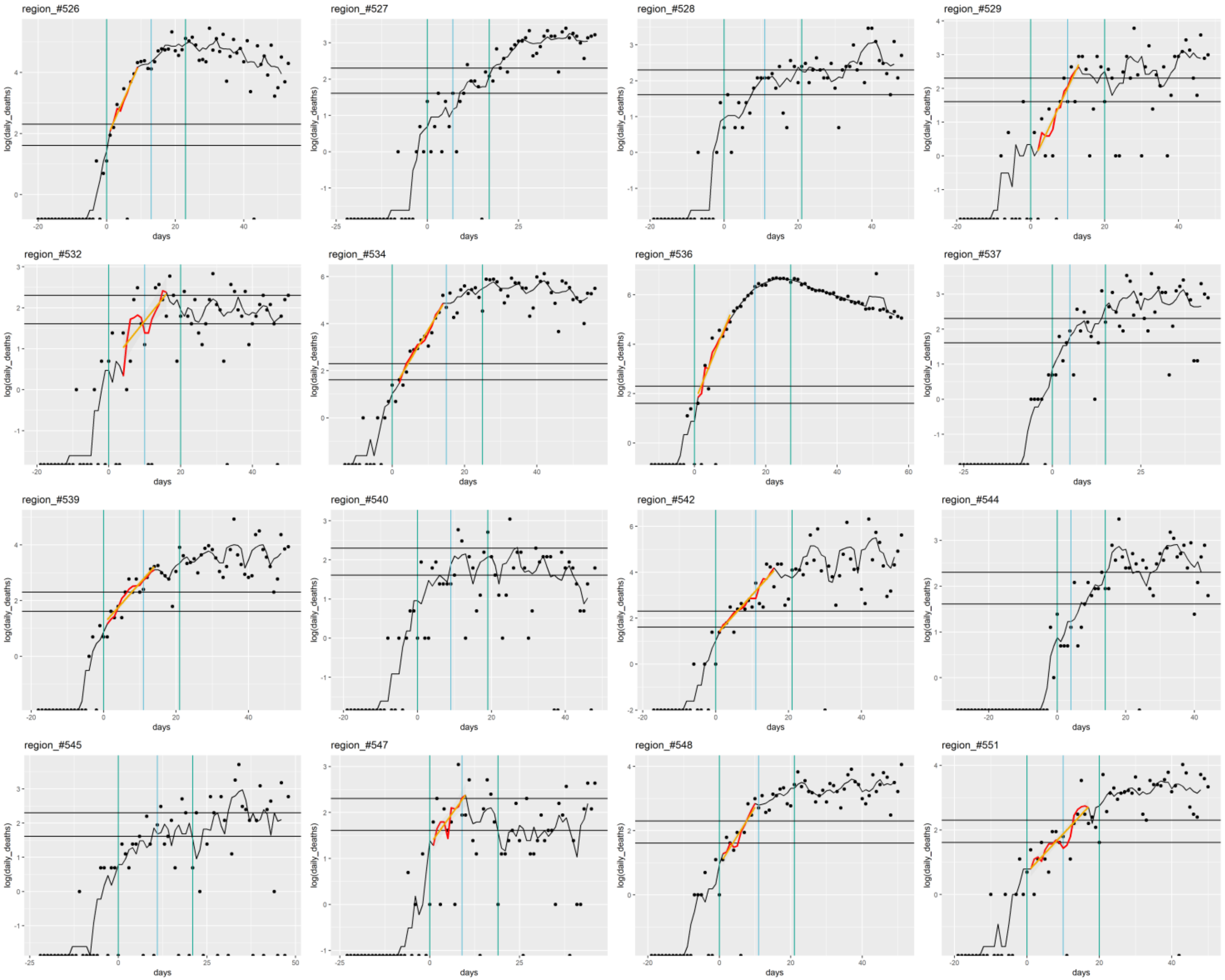

#### USA (continued) , Portugal (303-305) and Canada (624,635)

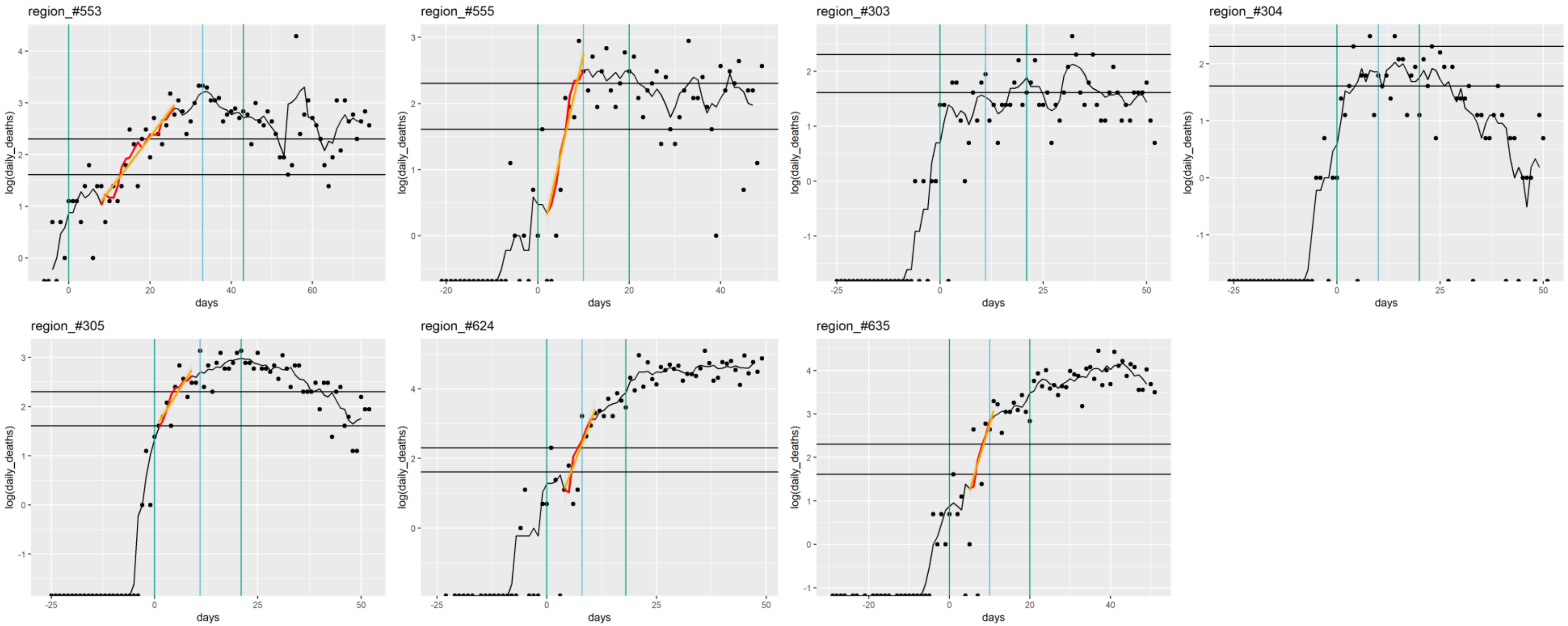

**Table S2: Results of the univariable hierarchical model assuming a linear relationship between covariates and R0 value**

| Variable (linear) | estimate | 95% confidence interval | p-value | AIC | Deviance explained (%) |
|---|---|---|---|---|---|
| Dew point temperature: min. | -0.075 | [-0.115 - -0.035] | 0.00049 | 146.22 | 19.02 |
| Dew point temperature: mean | -0.079 | [-0.122 - -0.037] | 0.00053 | 146.40 | 18.80 |
| Dew point temperature: max. | -0.081 | [-0.127 - -0.034] | 0.00123 | 148.00 | 16.55 |
| Absolute humidity: min. | -0.185 | [-0.295 - -0.074] | 0.00175 | 148.61 | 14.95 |
| Absolute humidity: mean | -0.162 | [-0.26 - -0.065] | 0.00187 | 148.74 | 14.77 |
| Absolute humidity: max. | -0.140 | [-0.23 - -0.051] | 0.00319 | 149.76 | 13.38 |
| Daily precip: mean | -0.092 | [-0.152 - -0.031] | 0.00411 | 150.24 | 12.72 |
| Temperature: min. | -0.075 | [-0.125 - -0.026] | 0.00432 | 150.51 | 14.30 |
| Temperature: mean | -0.072 | [-0.123 - -0.022] | 0.00653 | 151.47 | 15.09 |
| dist. to 1st region affected | -0.050 | [-0.089 - -0.01] | 0.01586 | 152.85 | 12.38 |
| Population density (log10) | 0.808 | [0.153 - 1.463] | 0.01868 | 153.05 | 8.74 |
| Temperature: max. | -0.056 | [-0.104 - -0.008] | 0.02606 | 154.11 | 12.28 |
| Population density | 0.001 | [0 - 0.002] | 0.05064 | 154.84 | 6.12 |
| Windspeed: mean | -0.035 | [-0.235 - 0.165] | 0.73087 | 158.69 | 0.20 |
| % pop over 80 | 0.021 | [-0.103 - 0.145] | 0.74377 | 158.68 | 1.17 |
| % pop over 70 | 0.011 | [-0.057 - 0.079] | 0.74549 | 158.70 | 0.63 |

**Table S3: Results of the univariable hierarchical model assuming a non-linear relationship between covariates and R0 value**

| Variables (B-spline) | p-value | AIC | GCV | Deviance explained (%) |
|---|---|---|---|---|
| DP: min. | 0.0004762 | 146.2165 | 0.5785073 | 19.01842 |
| DP: mean | 0.0005223 | 146.3991 | 0.5801903 | 18.79512 |
| Temp: mean | 0.0041478 | 146.4266 | 0.5845332 | 26.75646 |
| Daily precip: mean | 0.0173321 | 146.8934 | 0.5935806 | 30.99777 |
| dist. to 1st region affected | 0.0071834 | 146.8959 | 0.5859508 | 21.37092 |
| Absolute humidity: mean | 0.0042660 | 147.0754 | 0.5879088 | 21.75236 |
| Absolute humidity: min. | 0.0046919 | 147.3259 | 0.5902896 | 21.51877 |
| DP: max. | 0.0012101 | 148.0024 | 0.5951022 | 16.54749 |
| Temp: min. | 0.0074311 | 148.0998 | 0.5978279 | 21.02545 |
| Temp: max. | 0.0094238 | 148.5818 | 0.6049323 | 24.27944 |
| Absolute humidity: max. | 0.0081602 | 148.8980 | 0.6045329 | 18.04753 |
| % pop over 80 | 0.0248805 | 148.9918 | 0.6173410 | 31.38356 |
| % pop over 70 | 0.4606301 | 157.5315 | 0.6956170 | 10.40253 |
| Population density | 0.1300157 | 154.6511 | 0.6623683 | 10.28745 |
| Windspeed: mean | 0.4369166 | 157.7554 | 0.7003610 | 13.15879 |

Based on the results of table S2 and S3, the log10 of population density was included as a linear covariate, due to a small gain in AIC with a limited loss in deviance explained. Distance to the first region affected was included as a spline to avoid issues in log transformation of 0 values (for regions first affected). Weather/climate parameters were included as non-linear predictors.

**Table S4: Multivariable results for the relationship between R0 and weather parameters, under the assumption of a linear relationship obtained with the hierarchical generalized additive model.** Weather parameters are temperature, absolute humidity, and dew point temperature, adjusted for distance to the first region affected, population density, and elderly population. The linear approximation appears valid for dew point temperature: the spline had a linear shape (Figure 6) and the linear approximation does not modify the percentage of deviance explained. The linear approximation results in a minor decrease in deviance explained for absolute humidity. It appears however not relevant for temperature since it results in a strong decrease in deviance explained.

| Model | Variable | Estimate | 95%CI | p-value |
|---|---|---|---|---|
| **Model 1b** | Intercept | 1.48 | [-0.34 - 3.3] | 0.11618 |
| | Population density (log10) | 0.70 | [0.06 - 1.34] | 0.03562 |
| | % population over 80 | 0.04 | [-0.1 - 0.17] | 0.61838 |
| | Distance to first region affected in the country/coast | **spline** | | 0.0639 |
| | **Mean temperature** | -0.08 | [-0.13 - -0.02] | 0.01163 |
| | Dev. explained: 34.4% | | | |
| **Model 2b** | Intercept | 2.36 | [0.6 - 4.11] | 0.01081 |
| | Population density (log10) | 0.47 | [-0.14 - 1.08] | 0.13580 |
| | % population over 80 | 0.02 | [-0.1 - 0.15] | 0.72173 |
| | Distance to first region affected in the country/coast | **spline** | | 0.0654 |
| | **Mean AH** | -0.15 | [-0.26 - -0.03] | 0.01498 |
| | Dev. explained: 32.0% | | | |
| **Model 3b** | Intercept | 1.46 | [-0.2 - 3.13] | 0.09023 |
| | Population density (log10) | 0.49 | [-0.12 - 1.1] | 0.12005 |
| | % population over 80 | 0.05 | [-0.08 - 18] | 0.47088 |
| | Distance to first region affected in the country/coast | **spline** | | 0.0975 |
| | **Mean Dew Point Temperature** | -0.07 | [-0.12 - -0.02] | 0.00498 |
| | Dev. explained: 34.6% | | | |

**Figure S5: Sensitivity analysis of the link between weather covariates and R0 at different lags in a univariate model assuming a linear relationship.**

Lag=0 (green line) indicates the model presented in Figure 5, lag=-1 indicates a weather summary period 7 days later, lag=1 indicates a weather summary period 7 days earlier…

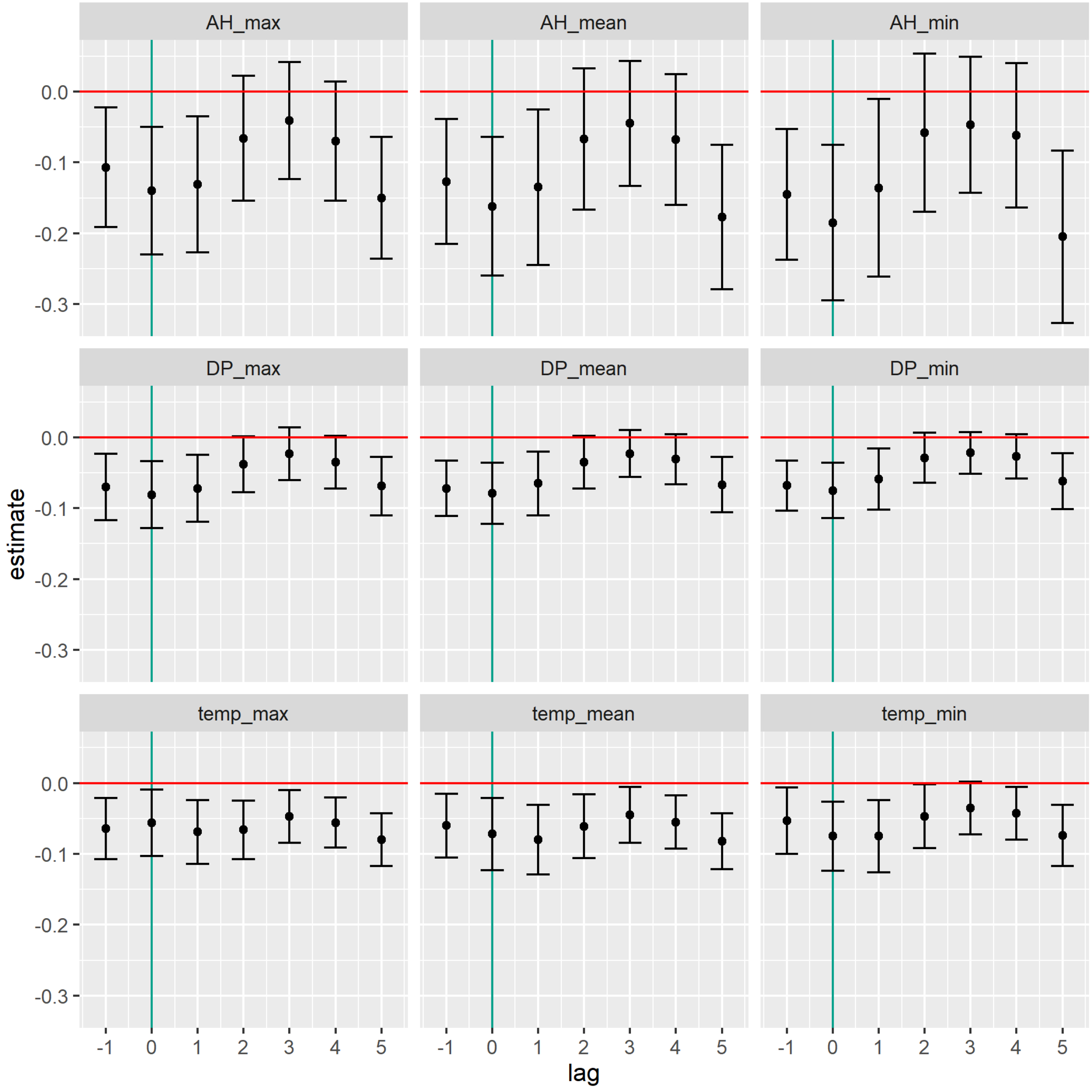

**Figure S6**

Sensitivity analysis for mean weather variable splines in the multivariate model.

Lag=0 indicates the final model, as shown in Figure 6, lag=-1 indicates a weather summary period 7 days later, lag=1 indicates a weather summary period 7 days earlier etc.

The effects of log10(population density) and percentage of population aged>80 were stable across lags.

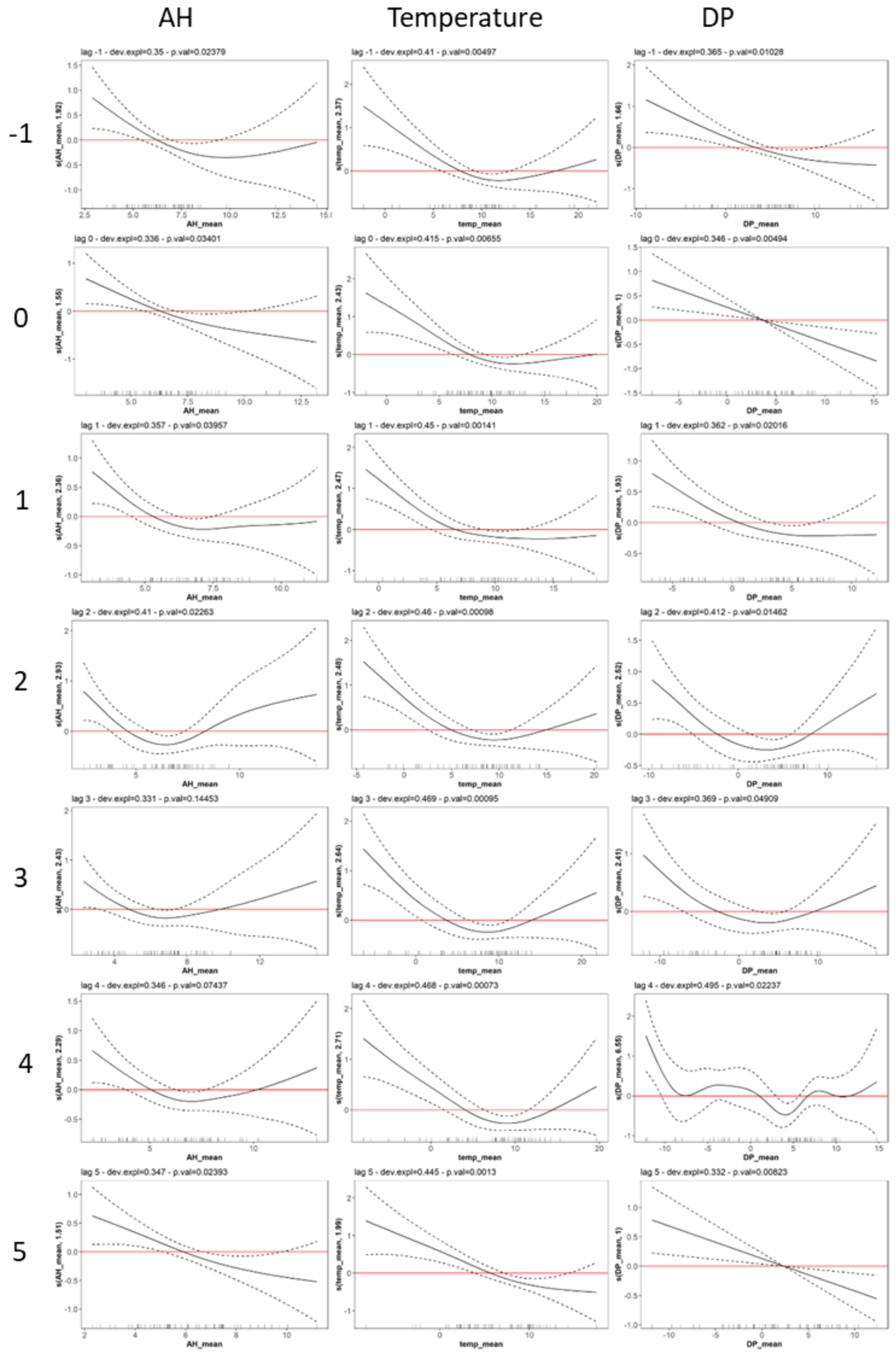

**Figure S7: Spearman correlation coefficient between lagged weather variables used in sensitivity analysis**

T: mean temperature, AH: mean absolute humidity, DP: mean dewpoint temperature

0: estimated transmission period (3-week lag from R0 calc. period); -1, 1-5: lagged observations

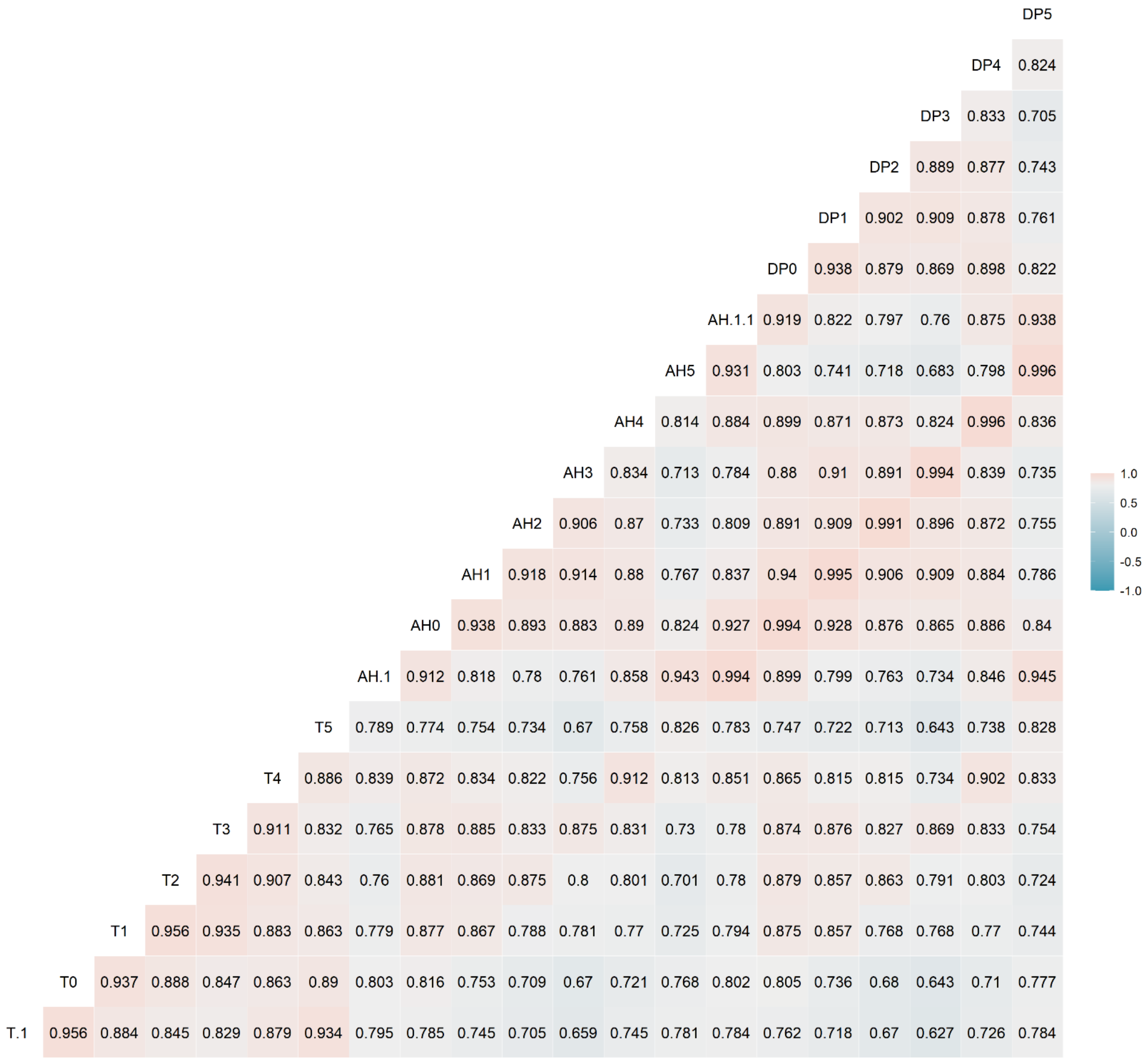

**Figure S8: Schematic representation of time periods used in the study**

The study period (green) was defined as the period where the growth of death counts was considered to occur without early stochasticity (cumulative death count>10) and with limited influence from lockdown (date≤lockdown+28 days).

Within the study period, the exponential growth period (in blue) was defined as the linear portion of the log(daily death count)=f(t) curve (black), and R0 was estimated on this period.

The transmission period (in red) was defined as the period when infections corresponding to the exponential growth period were acquired, i.e. 3 weeks earlier, over the same duration. Weather variable summary values were calculated over the transmission period.

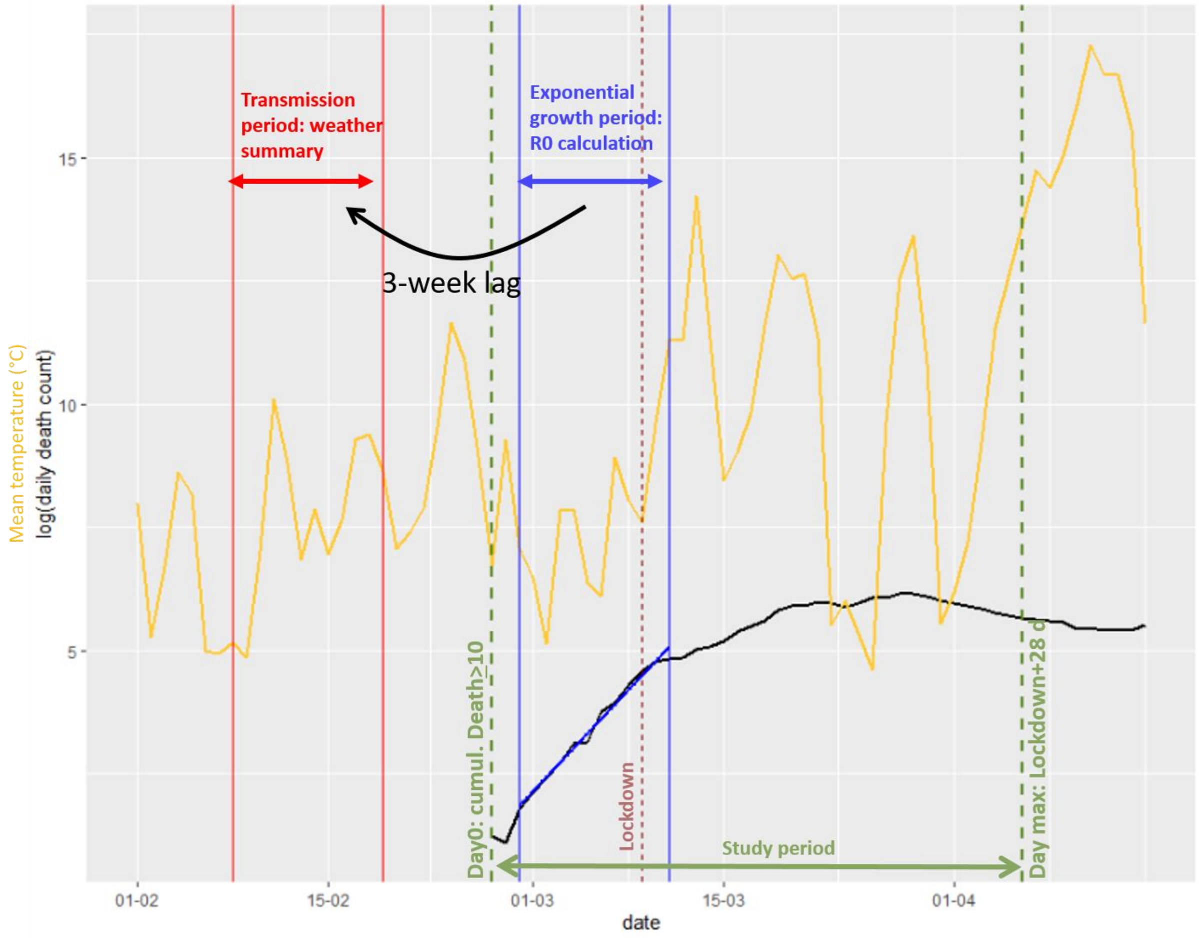
